# Anti-Ro52 autoantibodies modulate the transcriptomic effects of autoantibodies targeting intracellular autoantigens

**DOI:** 10.64898/2026.11.17.26368327

**Authors:** Mesra S. Ebadi, Jon Musai, Travis B. Kinder, Maria Casal-Dominguez, Stefania Dell’Orso, Shamima Islam, Gustaf Wigerblad, Gustavo Gutierrez-Cruz, Jose C. Milisenda, Albert Selva-O’Callaghan, Blake Warner, Janet E. Pope, Andreu Fernández-Codina, Peter D. Burbelo, Iago Pinal-Fernandez, Andrew L. Mammen

## Abstract

**Objective:** Ro52/TRIM21 is a cytoplasmic protein that binds intracellular antibodies and drives their proteasomal degradation. Autoantibodies targeting Ro52 frequently co-occur with myositis-specific autoantibodies, including anti-MDA5, and patients with both autoantibodies have greater disease severity and interstitial lung disease risk. We previously demonstrated that internalized anti-MDA5 autoantibodies promote interferon-stimulated gene expression in cultured myoblasts. Here, we investigated the interactions between anti-Ro52 and coexisting autoantibodies, including anti-MDA5.

**Methods:** Primary human skeletal muscle myoblasts were electroporated with purified IgG from healthy controls or patients with anti-MDA5, anti-Mi-2, or anti-PM/Scl antibodies, combined with anti-Ro52-positive IgG from patients with myositis, Sjögren’s disease, or systemic sclerosis. Affinity-purified anti-Ro52 was also used to validate findings obtained using total IgG. Cells underwent bulk RNA sequencing 24 hours after internalization.

**Results:** Co-internalization of anti-Ro52 with anti-MDA5 enhanced the anti-MDA5-induced type I interferon transcriptional response and expression of type II interferon-inducible genes compared with anti-MDA5 and control IgG. Increased expression of *IFNB1*, *ISG15*, *MX1*, and the interferon-γ-inducible inflammatory chemokines *CXCL9* and *CXCL10* was observed across independent patient-derived antibody combinations, anti-Ro52 preparations from multiple autoimmune diseases, and affinity-purified anti-Ro52. Conversely, co-internalization of anti-Ro52 with anti-Mi-2 or anti-PM/Scl reduced expression of transcripts associated with these nuclear autoantibodies.

**Conclusions:** These findings demonstrate that anti-Ro52 autoantibodies potentiate the activation of interferon pathways stimulated by the internalization of anti-MDA5 antibodies. We propose that in patients with anti-MDA5 autoantibodies, co-existing anti-Ro52 antibodies may exacerbate disease by inhibiting the proteasomal degradation of internalized anti-MDA5 antibodies and thereby enhancing the expression of interferon and interferon-stimulated genes.

## INTRODUCTION

Inflammatory myopathies (IMs) are a heterogeneous group of systemic autoimmune diseases affecting skeletal muscle, skin, lungs, and joints [1]. Myositis-specific autoantibodies identify disease subsets with characteristic clinical manifestations and prognoses [1]. Our analyses of muscle biopsies have further demonstrated that these subsets exhibit distinct transcriptional programs, including signatures consistent with dysfunction of the corresponding autoantigens [2–6]. These observations suggest that myositis autoantibodies contribute directly to disease biology beyond their established role as biomarkers.

Supporting this possibility, we and others previously showed that immunoglobulins accumulate within muscle fibers in compartments matching the localization of their target autoantigens, and that internalization of purified patient IgG into cultured human muscle cells reproduces abnormalities observed in patient biopsies [3, 7]. Subsequent spatial transcriptomics validated these signatures across independent cohorts and linked them to cellular injury and distinct inflammatory responses [2]. These findings support a pathogenic role for intracellular autoantibodies but leave unresolved how coexisting autoantibodies influence one another’s biological effects.

Anti-Ro52 is particularly relevant to this question because it frequently accompanies other myositis autoantibodies, especially anti-MDA5 and antisynthetase autoantibodies, both targeting cytoplasmic autoantigens [8–10]. Its presence has been associated with interstitial lung disease (ILD), and, in anti-MDA5-positive dermatomyositis, with rapidly progressive ILD and poorer outcomes [11–14]. These clinical associations raise the possibility that anti-Ro52 modifies the biological effects of coexisting cytoplasmic autoantibodies.

Ro52, encoded by *TRIM21*, is an E3 ubiquitin ligase and cytosolic Fc receptor involved in intracellular antibody recognition and innate immune regulation [15, 16]. TRIM21 can recognize the Fc region of intracellular antibodies and promote degradation of antibody-bound material. It also regulates interferon responses through ubiquitination of signaling proteins, including interferon regulatory factors [15]. These functions provide a rationale for investigating whether antibodies directed against Ro52 alter responses elicited by other intracellular autoantibodies.

Anti-MDA5 provides a clinically relevant model for examining these interactions. MDA5 is a cytoplasmic viral RNA sensor that promotes interferon production. In our previous experiments, intracellular delivery of IgG from anti-MDA5-positive patients induced *IFNB1* and interferon-stimulated gene expression in human myoblasts [3]. In contrast, anti-Mi-2 and anti-PM/Scl target nuclear complexes involved in transcriptional repression and RNA degradation, respectively [2, 3]. These autoantibodies therefore provide complementary models for determining whether anti-Ro52 modifies responses across distinct intracellular antigen contexts.

We hypothesized that intracellular anti-Ro52 modifies the transcriptional effects of coexisting autoantibodies. To test this hypothesis, we co-internalized primary human skeletal muscle myoblasts with patient-derived IgG containing anti-Ro52 and IgG containing anti-MDA5, anti-Mi-2, or anti-PM/Scl. By comparing their transcriptional responses, we sought to determine whether anti-Ro52 acts as a context-dependent modifier of autoantibody-associated cellular programs.

## METHODS

We used serum samples from patients enrolled in institutional review board-approved protocols at the National Institutes of Health, Western University, Vall d’Hebron University Hospital (Barcelona), and Hospital Clinic (Barcelona), and pooled serum from healthy donors at the NIH Clinical Center. Patient sample inclusion required seropositivity for one of the autoantibodies of interest. For myositis patients, fulfillment of NIH myositis criteria was also required [17]. Autoantibody status was confirmed using EUROLINE Autoimmune Inflammatory Myopathies Profile and Luciferase Immunoprecipitation System (LIPS) assay as previously reported [18–22].

### Standard protocol approvals and patient consent

This study was approved by the Institutional Review Boards of the National Institutes of Health, Western University, Vall d’Hebron University Hospital (Barcelona), and Hospital Clinic (Barcelona). Written informed consent was obtained from each participant. All methods were performed in accordance with the relevant guidelines and regulations.

### Electroporation

Human immunoglobulin G (IgG) from serum was purified using Protein G Agarose (Millipore, ref. 16-266) and the Amicon Pro Purification Device (Millipore Sigma, ref. ACS500024) and concentrated using the Amicon Ultra Centrifugal Filter, 30-kDa molecular-weight cutoff (Millipore Sigma, ref. UFC503024).

Normal human skeletal muscle myoblasts (Lonza, CC-2580) cultured in growth medium (Lonza, CC-3245) were electroporated using the Lonza 4D-Nucleofector 96-well unit with program EN-150 with 50% purified IgG and 50% P5 Primary Cell 4D-Nucleofector X Kit L (Lonza, ref. V4XP-5024) at 100 µg for the first two experiments and 200 µg for subsequent experiments. After electroporation, cells were plated in differentiation medium and harvested for RNA extraction and subsequent RNA sequencing 24 hours later.

### RNA extraction, library preparation, and sequencing

RNA was isolated using the Qiagen RNeasy kits (ref. 74104 and 74181). Libraries were either prepared with the NEBNext Poly(A) mRNA Magnetic Isolation Module (New England BioLabs, ref. #E7490) and NEBNext Ultra II Directional RNA Library Prep Kit for Illumina (New England BioLabs, ref. #E7760), or the NEBNext UltraExpress RNA Library Prep Kit (New England BioLabs, ref. #E3330L).

### Affinity purification

For affinity purification of anti-Ro52 autoantibodies, 60 μg of full-length, wild-type Ro52 protein (ProSpec, catalog no. PRO-328) was covalently coupled to 300 μL of Pierce NHS-Activated Magnetic Beads (Thermo Fisher Scientific, catalog no. 88826). The beads were incubated for 1 h with 1 mL of purified IgG (1 mg/mL) from an anti-Jo-1–positive, anti-Ro52–positive patient (patient 60). Bound antibodies were eluted with 600 μL of 0.1 M glycine (pH 2.5) and neutralized with 1 M Tris (pH 8.5). Affinity purification was repeated three times using a single patient sample, and the eluates were pooled and concentrated using centrifugal filters as described below. Anti-Ro52 reactivity was confirmed by line blot, and IgG concentration was measured by human IgG ELISA. Total IgG (50 μg) from an anti-MDA5–positive patient (sample MDA5-1) or healthy controls was co-internalized with 2 μg of affinity-purified anti-Ro52 or healthy control IgG. RNA expression was assessed as described above.

For affinity purification of antibodies recognizing the N-terminal region of Ro52, Escherichia coli was transformed with a plasmid encoding Ro52 residues 2–276 and cultured in Luria–Bertani (LB) medium containing the appropriate antibiotics. The recombinant fragment was purified using agarose resin and used to affinity-purify anti-Ro52 autoantibodies from the serum of a patient with Sjögren’s syndrome. Antibody purification was performed using gravity-flow columns, with glycine used for elution. Anti-Ro52 reactivity was confirmed by LIPS.

### Interferon treatment of human skeletal muscle myoblasts

Normal human skeletal muscle myoblasts were cultured according to the protocol recommended by the supplier (Lonza). When 80% confluent, myoblasts were induced to differentiate into myotubes by replacing the growth medium with differentiation medium (DMEM-F12 [Lonza, ref. 12-719F] supplemented with 2% horse serum [Gibco, ref. 16050-122], insulin-transferrin-selenium [Gibco, ref. 41400-045], and penicillin-streptomycin-L-glutamine [Gibco, ref. 10378-016]). Cells were harvested before differentiation and then daily after differentiation for 6 days.

To examine the effect of different types of interferon on gene expression we treated the cells daily with 1000 U/mL of IFN-β (PeproTech, ref. 300-02BC) and 1000 U/mL of IFN-γ (PeproTech, ref. 300-02), respectively, for 7 days. Then, the cells were harvested for RNA extraction and RNA sequencing.

### Quantification of *MX1* expression by TaqMan RT-qPCR

Twenty-four hours after internalization, cells were harvested and analyzed using the Cells-to-CT 1-Step TaqMan Kit (Thermo Fisher Scientific, A25603) according to the manufacturer’s protocol. TaqMan assays were performed on a QuantStudio 12K Flex instrument running software v1.6, with *18S* rRNA (Hs99999901_s1, VIC-MGB_PL) multiplexed with *MX1* (Hs00895608_m1, FAM-MGB).

### Statistics

For RNA-seq, reads were aligned to GRCh38-based reference annotations and processed using previously reported pipelines [4, 23–27]. Raw counts were normalized with the trimmed mean of M values method, and differential expression was assessed with limma/edgeR. ReactomePA was used for pathway analysis. P values were adjusted by the Benjamini-Hochberg method, with q<0.05 considered significant.

IFN-β- and IFN-γ-predominant genes were defined in an independent human skeletal muscle myoblast dataset using three unadjusted limma-voom comparisons: each IFN at 1,000 units versus untreated and IFN-β versus IFN-γ. Predominant genes were significantly upregulated versus untreated and the other IFN (Benjamini–Hochberg-adjusted P <0.05 for both comparisons). Gene lists were ranked by ascending raw P value from the direct IFN-β versus IFN-γ comparison, following limma output; results for genes shown in the figures are provided in Supplementary Table S1.

## RESULTS

### Co-internalization of anti-Ro52 antibodies from patients with the antisynthetase syndrome enhances anti-MDA5 antibody-induced IFN gene expression in cultured myoblasts

To assess the transcriptomic effects of anti-Ro52 autoantibodies obtained from multiple patients, HSMM cells were internalized with pairwise combinations of anti-Ro52 IgG from three antisynthetase syndrome patients and anti-MDA5 IgG from three dermatomyositis patients without anti-Ro52 antibodies.

Internalization of anti-Ro52 IgG alone had minimal effects on gene expression. As previously observed [3], internalization of anti-MDA5 IgG from all three patients induced both a robust type I and type II interferon-inducible gene signature. Co-internalization of anti-Ro52 and anti-MDA5 IgG increased expression of both type I and type II pathway genes compared with internalization of anti-MDA5 IgG plus control. Specifically, expression of *IFNB1, ISG15,* and *MX1* was increased following co-internalization. Co-internalization also increased expression of IFNG-inducible inflammatory chemokines *CXCL9* and *CXCL10*, whereas *IFNG* expression remained unchanged. Increased expression of IFN pathway genes and inflammatory chemokines was consistently observed across the paired anti-Ro52/anti-MDA5 donor combinations (Figure 1A; Figures S1 and S2).

**Figure 1.**
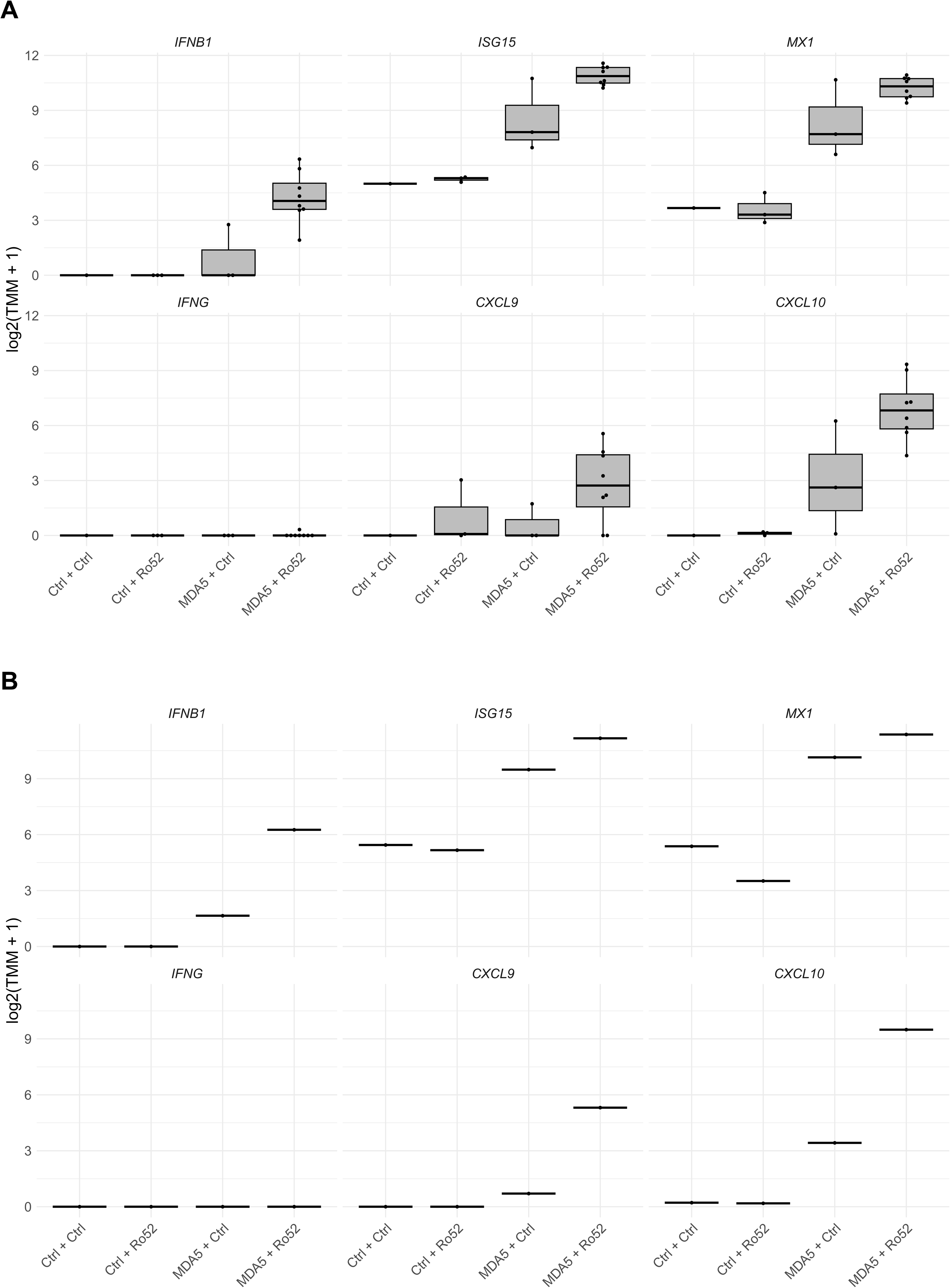
Co-internalization of anti-Ro52 and anti-MDA5 autoantibodies enhances type I and type II gene expression. **(A)** Primary human skeletal muscle myoblasts were internalized with combinations of healthy control Ig, anti-Ro52-positive IgG (from n = 3 patients), and anti-MDA5-positive IgG (from n = 3 patients). **(B)** A separate validation experiment using one anti-Ro52/anti-MDA5 IgG pair.

To confirm these findings, we repeated the experiment using one anti-Ro52 IgG/anti-MDA5 IgG pair under identical conditions. Similar to the initial experiment, co-internalization of anti-Ro52 and anti-MDA5 IgG increased expression of both type I and type II IFN-inducible genes. Whereas IFNB1 was overexpressed, *IFNG* expression remained unchanged (Figure 1B; Figures S3 and S4).

To determine whether affinity-purified anti-Ro52 antibodies recapitulated the effects observed with whole IgG preparations, affinity-purified anti-Ro52 antibody was co-internalized with total anti-MDA5-positive IgG. Affinity-purified anti-Ro52 antibodies replicated the enhancement of an IFN signature observed following co-internalization with whole anti-Ro52 IgG (Figures 2 and 3; Figures S5 and S6; Supplementary Table S2). These data illustrate that the enhancement of the anti-MDA5-induced IFN response was due to the Ro52-specific antibodies and not other antibodies in the whole IgG preparations.

**Figure 2.**
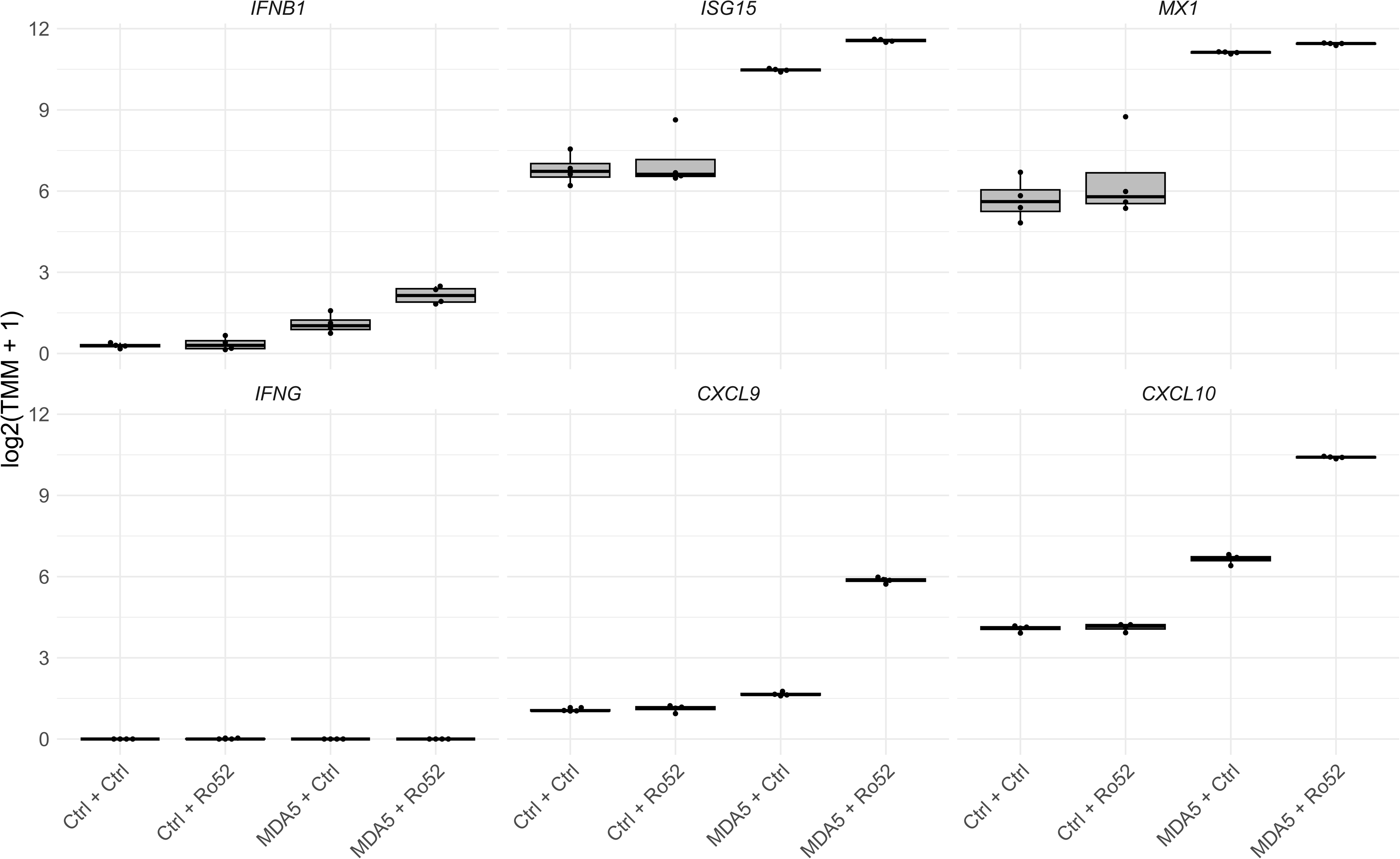
Affinity-purified anti-Ro52 enhances the anti-MDA5-associated IFN transcriptional response. Primary human skeletal muscle myoblasts were co-internalized with healthy-control or anti-MDA5-positive IgG and control IgG or anti-Ro52 affinity-purified using full-length Ro52.

**Figure 3.**
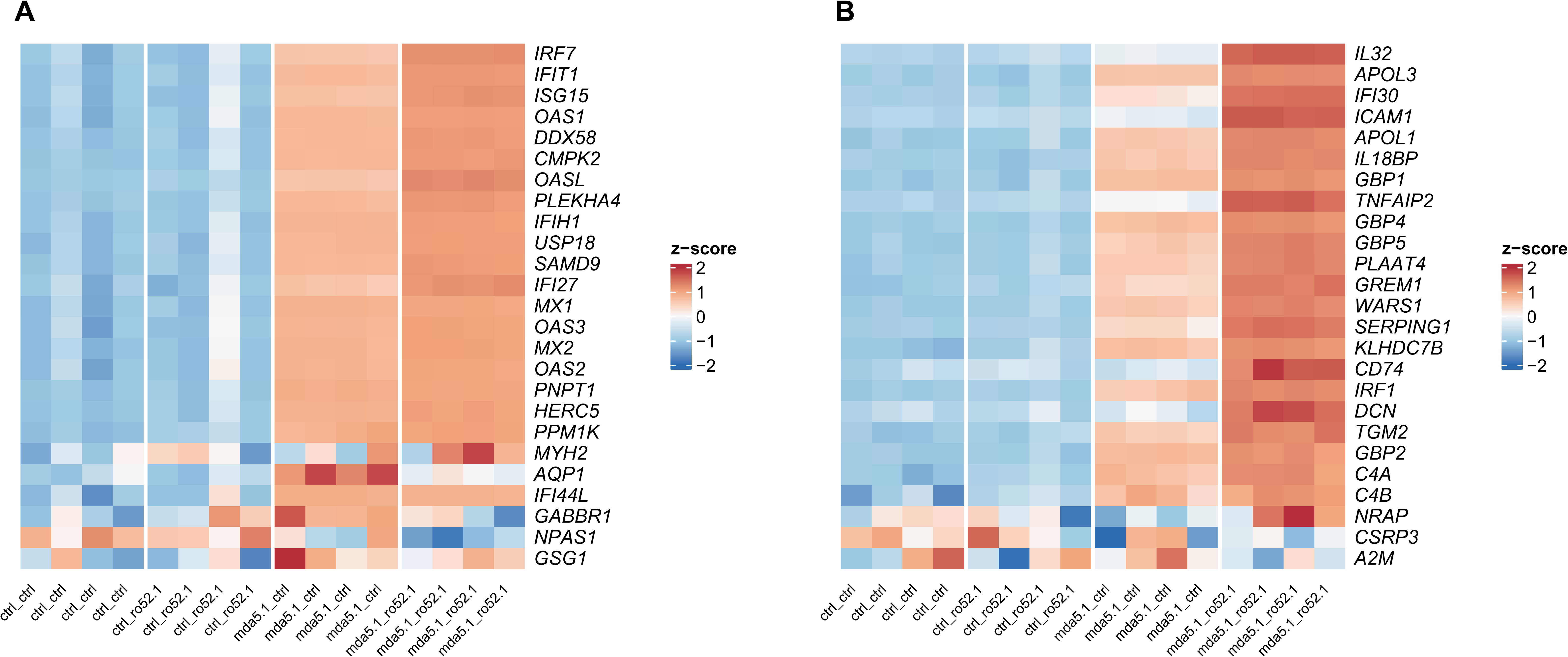
IFN-predominant transcriptional responses following co-internalization of affinity-purified anti-Ro52 and anti-MDA5 IgG. Primary human skeletal muscle myoblasts were co-internalized with the indicated patient-derived or healthy-control IgG preparations, and RNA was collected 24 hours later. Healthy-control or anti-MDA5-positive IgG was combined with control IgG or anti-Ro52 antibodies affinity-purified using full-length Ro52. Four RNA-seq libraries are shown per condition, for 16 libraries in total. (A) The first 25 IFN-beta-predominant genes, including *MX1* and *ISG15*. (B) The first 25 IFN-gamma-predominant genes.

Interestingly, internalization of anti-Ro52 autoantibodies recognizing the N-terminal region of Ro52, which contains the E3 ubiquitin ligase domain but not the immunoglobulin-binding domain, did not reproduce the enhancement of *MX1* expression observed following co-internalization with whole anti-Ro52-positive IgG (Figure S7), suggesting that active anti-Ro52 autoantibodies are those blocking the immunoglobulin Fc-binding region of Ro52.

### Co-internalization of anti-Ro52 antibodies from patients with Sjogren’s disease or systemic sclerosis also augments anti-MDA5-induced IFN gene expression in cultured myoblasts

To determine whether the disease origin of anti-Ro52 autoantibodies influences biological activity, anti-Ro52 IgG from patients with myositis (anti-Ro52.1), Sjögren’s disease (anti-Ro52.4), or systemic sclerosis (anti-Ro52.5) was independently co-internalized with anti-MDA5 IgG into HSMM cells. Co-internalization of anti-MDA5 IgG with anti-Ro52 IgG from each disease source increased expression of type I IFN genes, including *IFNB1*, *ISG15*, and *MX1*, compared with anti-MDA5 IgG plus control. Similarly, expression of the IFNG-inducible chemokines *CXCL9* and *CXCL10* was increased, whereas *IFNG* expression remained unchanged (Figures 4 and 5; Figures S8 and S9; Supplementary Table S3).

**Figure 4.**
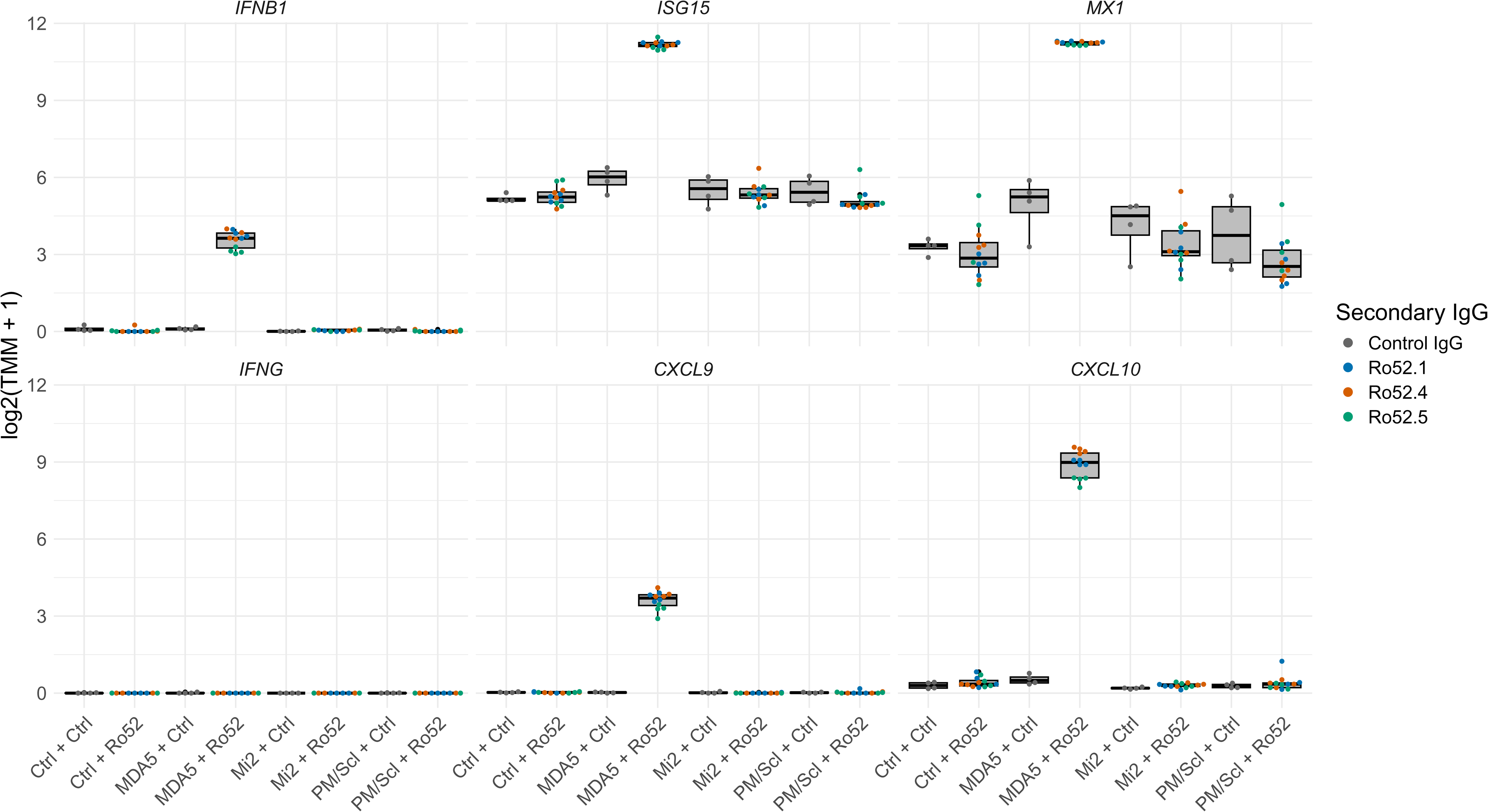
Anti-Ro52 autoantibodies from multiple autoimmune diseases augment anti-MDA5-induced interferon-β pathway gene expression. Primary human skeletal muscle myoblasts were co-internalized with control, anti-MDA5-positive, anti-Mi-2-positive, or anti-PM/Scl-positive IgG and control IgG or anti-Ro52-positive IgG from patients with myositis (Ro52.1, blue), Sjögren’s disease (Ro52.4, brown), or systemic sclerosis (Ro52.5, green). RNA was profiled 24 hours after internalization.

**Figure 5.**
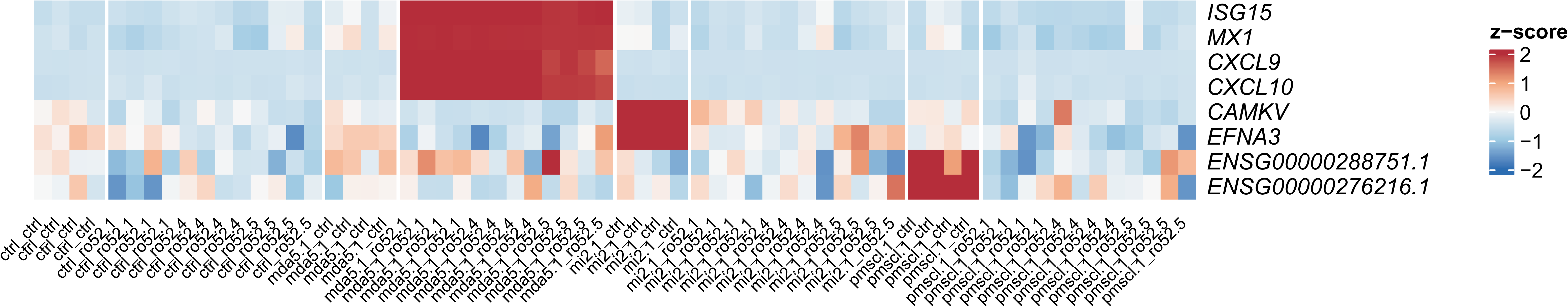
Representative transcriptional responses to anti-Ro52 co-internalization across autoantibody specificities. Primary human skeletal muscle myoblasts were co-internalized with patient-derived IgG containing anti-MDA5, anti-Mi-2, or anti-PM/Scl, or with healthy-control IgG, each combined with either control IgG or anti-Ro52-positive IgG from patients with myositis (Ro52.1), Sjögren disease (Ro52.4), or systemic sclerosis (Ro52.5). RNA was collected 24 hours after internalization. *ISG15* and *MX1* are representative *IFNB1*-inducible genes; *CXCL9* and *CXCL10* are IFNG-inducible genes; *CAMKV* and *EFNA3* represent Mi-2-specific genes; and the two transcripts labeled by Ensembl identifiers represent PM/Scl-specific genes.

Gene set enrichment analysis demonstrated marked enrichment of canonical type I and type II IFN signaling pathways, which clustered together. In contrast, pathways involved in mitosis and cell-cycle progression were negatively enriched, consistent with a shift toward an antiviral transcriptional program (Figure 6).

**Figure 6.**
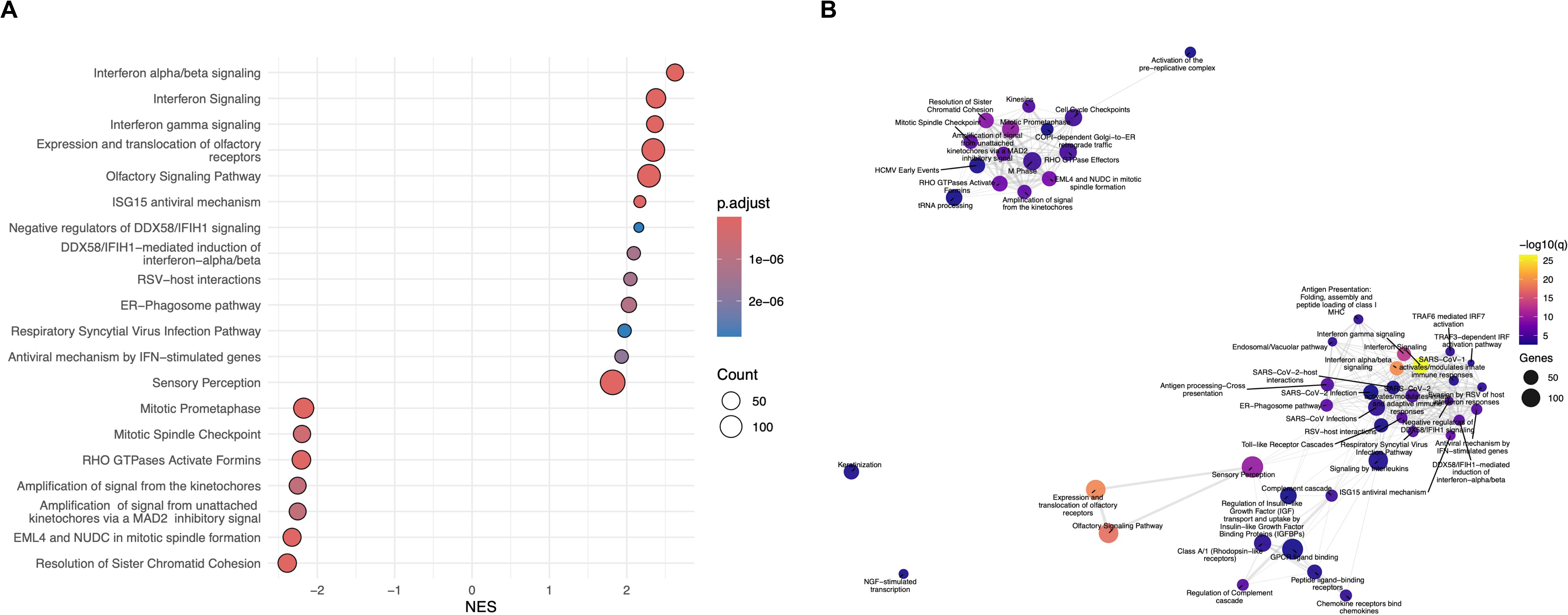
Gene set enrichment and enrichment network analyses demonstrate activation of coordinated type I IFN signaling following co-internalization of anti-Ro52 and anti-MDA5 autoantibodies. **(A)** Gene set enrichment analysis (GSEA) following co-internalization of anti-Ro52-positive and anti-MDA5-positive IgG. Pathways are plotted by normalized enrichment score (NES). Positive NES indicates enrichment toward anti-MDA5 plus anti-Ro52 relative to anti-MDA5 plus control IgG; negative NES indicates enrichment in the opposite direction. Dot size represents the contributing gene count, and dot color indicates the adjusted *P* value. **(B)** Enrichment network of pathways returned by GSEA. Each node represents a pathway; node size indicates the contributing gene count and node color indicates enrichment significance (-log10 adjusted P value). Edges connect pathways with overlapping contributing genes.

### Anti-Ro52 co-internalization decreases the expression of the anti-Mi2-specific and anti-PM/Scl-specific gene sets

To determine whether anti-Ro52 modulates transcriptional responses induced by nuclear autoantibodies, anti-Ro52 IgG was independently co-internalized with anti-Mi-2 IgG or anti-PM/Scl IgG into HSMM. Co-internalization with anti-Ro52 Ig reduced expression of the Mi-2-associated transcripts, such as *CAMKV* and *EFNA3,* compared with anti-Mi-2 IgG co-internalized with control IgG. Similarly, expression of the PM/Scl-associated long non-coding transcripts, such as *ENSG00000288751.1* and *ENSG00000276216.1,* was reduced following co-internalization of anti-Ro52 Ig with anti-PM/Scl IgG compared with anti-PM/Scl IgG co-internalized with control IgG (Supplementary Figure S10). In contrast to the enhanced type I IFN pathway gene expression observed with anti-MDA5, co-internalization of anti-Ro52 with nuclear autoantibodies reduced expression of Mi-2- and PM/Scl-associated transcripts.

To confirm these findings, the experiment was repeated under identical experimental conditions using anti-Ro52 IgG from patients with myositis, Sjögren’s disease, and systemic sclerosis. Similar to the initial experiment, co-internalization of anti-Ro52 Ig with anti-Mi-2 Ig or anti-PM/Scl IgG reduced expression of the Mi-2-associated transcripts *CAMKV* and *EFNA3 and* the PM/Scl-associated transcripts *ENSG00000288751.1* and *ENSG00000276216.1* (Figures 5 and 7; Figure S11; Supplementary Table S3).

**Figure 7.**
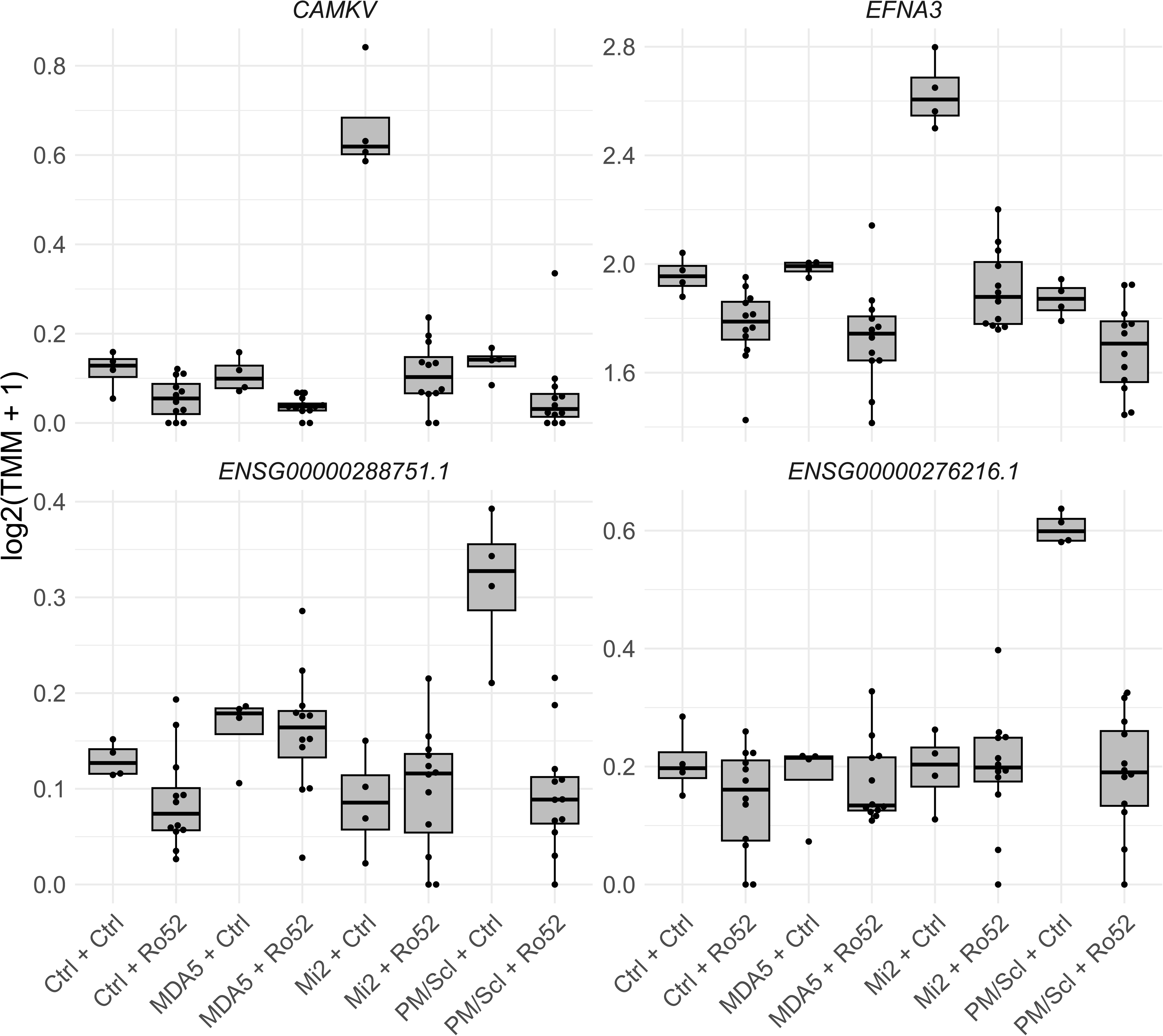
Independent replication of anti-Ro52-mediated modulation of Mi-2- and PM/Scl-associated transcripts. Primary human skeletal muscle myoblasts were co-internalized with control, anti-MDA5-positive, anti-Mi-2-positive, or anti-PM/Scl-positive IgG and control IgG or anti-Ro52-positive IgG from patients with myositis (Ro52.1), Sjögren’s disease (Ro52.4), or systemic sclerosis (Ro52.5). RNA was profiled 24 hours after internalization.

## DISCUSSION

In this study, we have shown that co-internalized anti-Ro52 antibodies modulate the intensity of transcriptional responses elicited by the internalization of myositis autoantibodies. Specifically, anti-Ro52 Ig amplified the IFN response induced by the internalization of anti-MDA5 Ig. Conversely, anti-Ro52 Ig attenuated the expression of transcripts uniquely associated with anti-Mi-2 Ig and anti-PM/Scl Ig internalization. These effects were reproduced using anti-Ro52 IgG from patients with myositis, Sjögren’s disease, and systemic sclerosis. Affinity-purified anti-Ro52 Ig also enhanced the anti-MDA5-induced IFN response, demonstrating that the observed effects were due to anti-Ro52 autoantibodies rather than other antibodies present in whole Ig fractions. Together, these findings identify anti-Ro52 antibodies as a functional modifier of the transcriptomic effects of myositis autoantibody internalization.

The interaction between anti-Ro52 and anti-MDA5 is particularly relevant given that their coexistence is associated with more severe disease, especially interstitial lung disease [11–14]. Although anti-MDA5 dermatomyositis is characterized by marked activation of IFN signaling [3, 24], the contribution of anti-Ro52 to this response has remained unclear [24]. Our findings demonstrate that intracellular anti-Ro52 amplifies an MDA5-driven IFN transcriptional program without independently inducing IFN signaling. Consistent with this effect, pathway analysis revealed enrichment of both type I and type II IFN–responsive gene sets. Notably, *IFNB1*, but not *IFNG*, was detectably upregulated, suggesting that enrichment of type II IFN–responsive genes may reflect downstream transcriptional effects of MDA5-associated signaling rather than increased *IFNG* expression. Together, these findings support a role for anti-Ro52 as a context-dependent amplifier of MDA5-associated IFN responses.

A notable finding was that the effects of anti-Ro52 depended on the accompanying autoantibody. Anti-Ro52 enhanced the interferon response induced by anti-MDA5 but attenuated the transcriptional changes associated with anti-Mi-2 and anti-PM/Scl. These findings suggest that anti-Ro52 modulates distinct transcriptional programs in a context-dependent manner rather than uniformly amplifying intracellular immune signaling. The contrasting effects observed with autoantibodies targeting cytoplasmic versus nuclear antigens raise the possibility that antigen localization contributes to this variability. Whether these effects help explain the co-occurrence of anti-Ro52 with anti-Jo-1 in myositis or with anti-Ro60 and anti-La in Sjögren’s disease remains to be established.

Interestingly, antibodies affinity-purified against full-length Ro52 enhanced the response to anti-MDA5, whereas those affinity-purified against the N-terminal region of Ro52 did not. The N-terminal region contains the E3 ubiquitin ligase domain but lacks the immunoglobulin Fc-binding domain. This suggests that the immunoglobulin-binding region may mediate the interaction between anti-Ro52 and anti-MDA5. One attractive mechanistic explanation is that the binding of anti-Ro52 autoantibodies to this region prevents the Ro52 protein from binding and subsequently degrading other immunoglobulins. This could allow more intracellular anti-MDA5 autoantibodies to accumulate and thereby enhance the IFN response driven by anti-MDA5 antibodies binding to MDA5 protein (Figure 8). Previous studies have suggested that anti-Ro52 autoantibodies inhibit the enzymatic activity of Ro52 [16], but their effects on its Fc-binding region have not been explored in depth.

**Figure 8.**
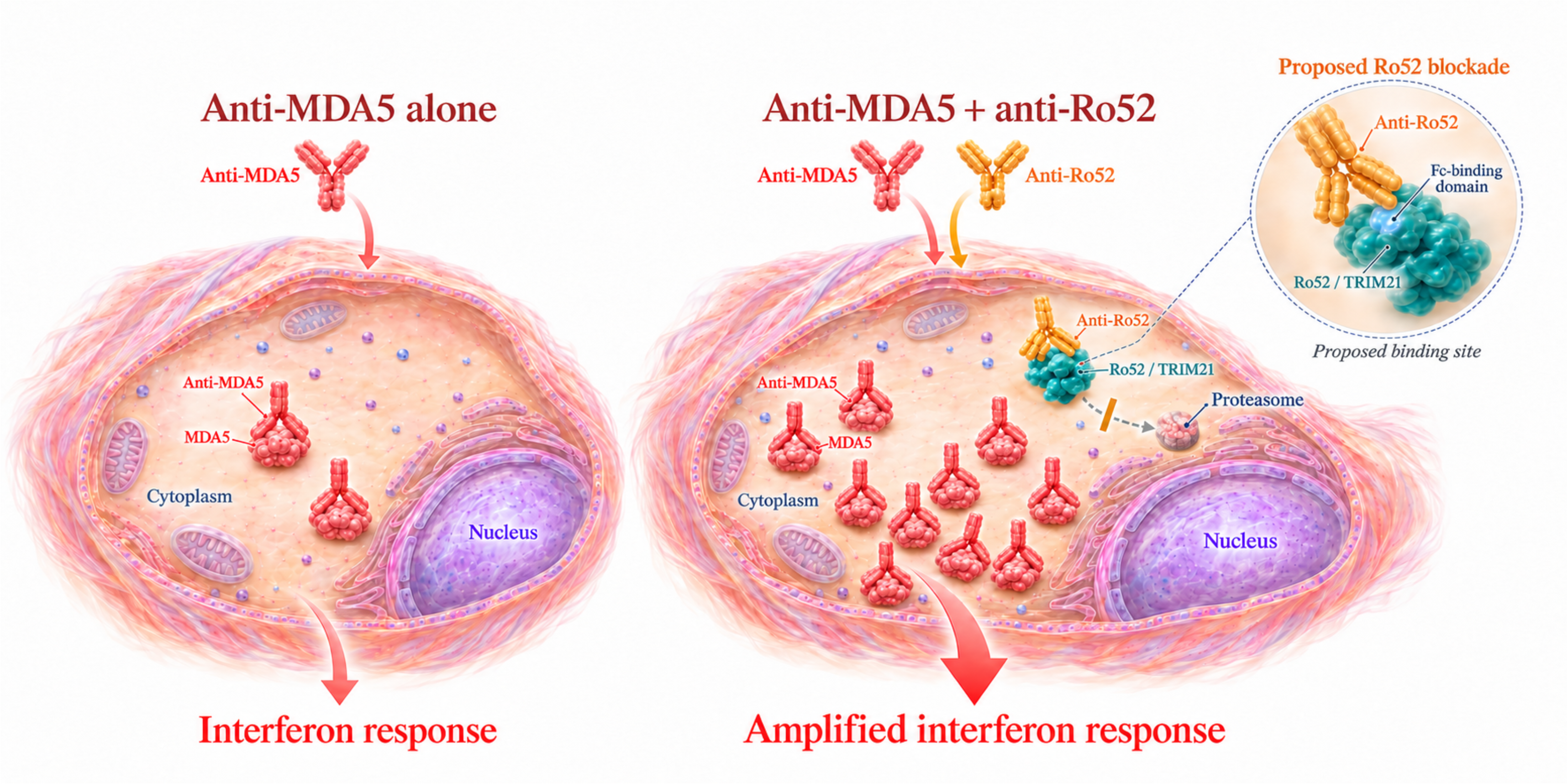
Proposed mechanism by which anti-Ro52 antibodies amplify the anti-MDA5-associated interferon response. Following experimental intracellular delivery, anti-MDA5 antibodies activate cytoplasmic MDA5 and induce interferon signaling. Anti-Ro52 antibodies impair Ro52/TRIM21 recognition and clearance of MDA5–anti-MDA5 immune complexes, prolonging their signaling and amplifying the interferon response.

Several limitations should be acknowledged. First, only a small number of patient-derived antibody samples were tested, although the principal findings were reproduced across independent experiments and with anti-Ro52 antibodies obtained from multiple autoimmune diseases. Second, electroporation provides an experimental model of intracellular antibody delivery and may not fully reflect physiological mechanisms of antibody internalization. Nevertheless, accumulating evidence supports the concept that myositis autoantibodies can gain intracellular access and exert functional biological effects [5]. Third, our study was limited to the interaction of anti-Ro52 with three autoantibodies, anti-MDA5, anti-Mi-2, and anti-PM/Scl. These findings cannot yet be generalized to all cytoplasmic or nuclear autoantibodies, and future studies of anti-Ro52 interactions with other autoantibodies are warranted. Finally, our primary objective was to determine whether anti-Ro52 modifies the effects of other autoantibodies, rather than to define the underlying mechanisms. These mechanisms are likely complex and require investigation of the balance between autoantigen synthesis and degradation.

In conclusion, our findings show that anti-Ro52 autoantibodies can modulate transcriptional responses elicited by myositis autoantibodies internalized into skeletal muscle cells. This suggests that anti-Ro52 antibodies may play a functional role in modifying disease severity rather than being simply a biomarker. By enhancing MDA5-associated interferon responses while attenuating those induced by anti-PM/Scl and anti-Mi-2, anti-Ro52 appears to act as a context-dependent regulator of intracellular immune signaling. These findings advance our understanding of anti-Ro52 function in myositis and suggest that interactions among coexisting autoantibodies may contribute to its molecular heterogeneity, with potential implications for other autoimmune diseases associated with intracellular autoantibodies.

## Contributorship

All authors contributed to study conception and design, data interpretation, substantive manuscript review, and approval of the final draft.

## Patient and public involvement

Patients and/or the public were not involved in the design, conduct, reporting, or dissemination plans of this research.

## Declaration of generative AI and AI-assisted technologies in the writing process

During the preparation of this work, the authors utilized OpenAI Codex to assist with drafting, code development, and figure preparation from experimental data. Following its use, the authors thoroughly reviewed and edited the content as necessary, taking full responsibility for the final version of the publication.

## Data Availability

All data produced in the present study are available upon reasonable request to the authors

