## Supplementary Figures for "Anti-Ro52 autoantibodies modulate the transcriptomic effects of autoantibodies targeting intracellular autoantigens"

**Supplementary Figure S1. Donor-detail plots for the initial MDA5 and validation experiments.** (A) Human skeletal muscle myoblasts were co-electroporated with IgG from three anti-MDA5-positive and three anti-Ro52-positive donors. (B) A separate validation used one anti-MDA5/anti-Ro52 donor pair and one library per condition. RNA was collected 24 hours later. Expression is log2(TMM-normalized counts per million + 1). Boxes show medians and interquartile ranges; points are individual RNA-seq libraries. Colors identify donor combinations; lines link samples sharing the primary IgG donor.

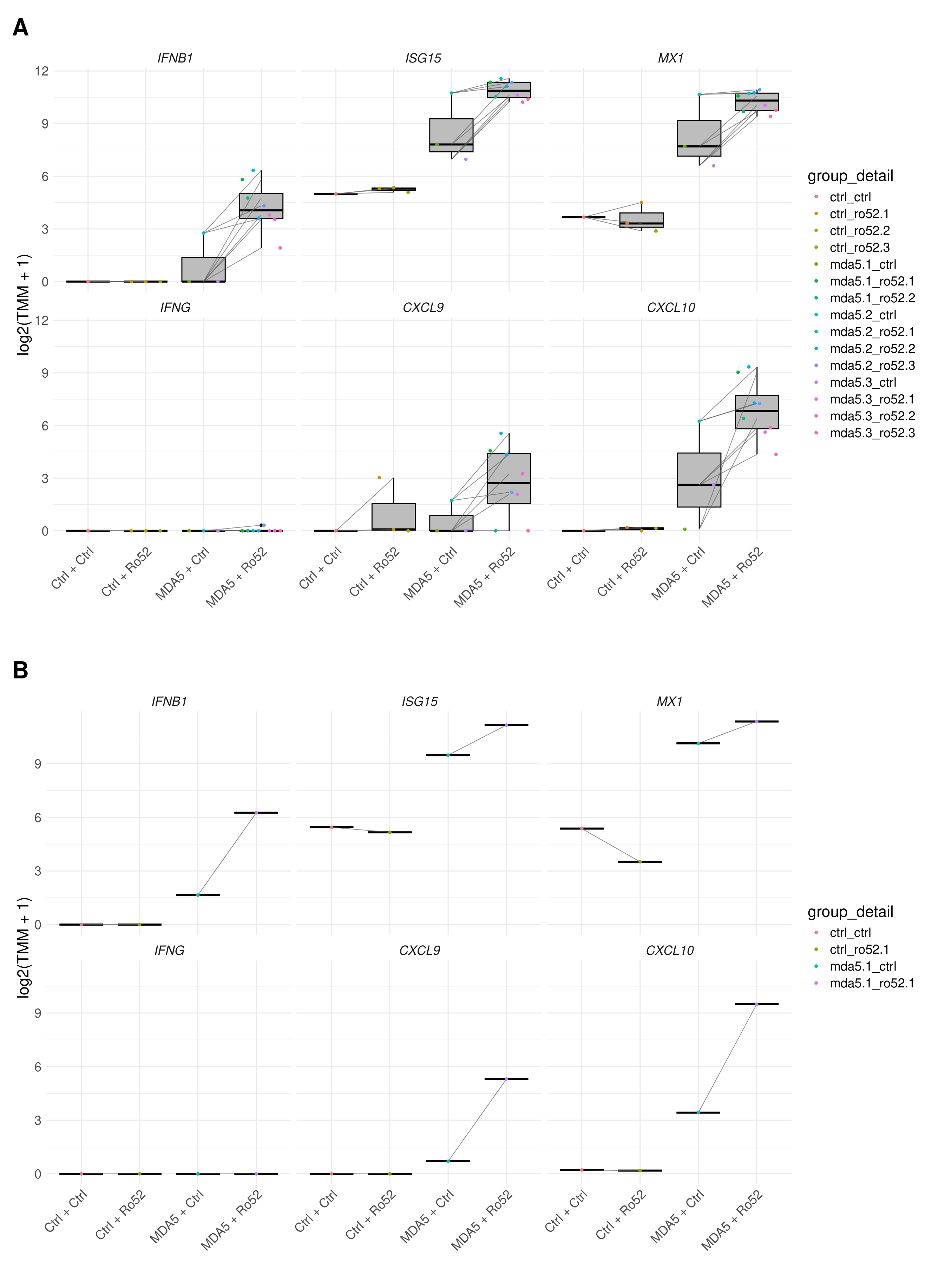

**Supplementary Figure S2. Representative-gene heatmap for the initial MDA5 experiment.** *ISG15*, *MX1*, *CXCL9*, and *CXCL10* expression in the 15 RNA-seq libraries from the initial experiment in Figure 1A.

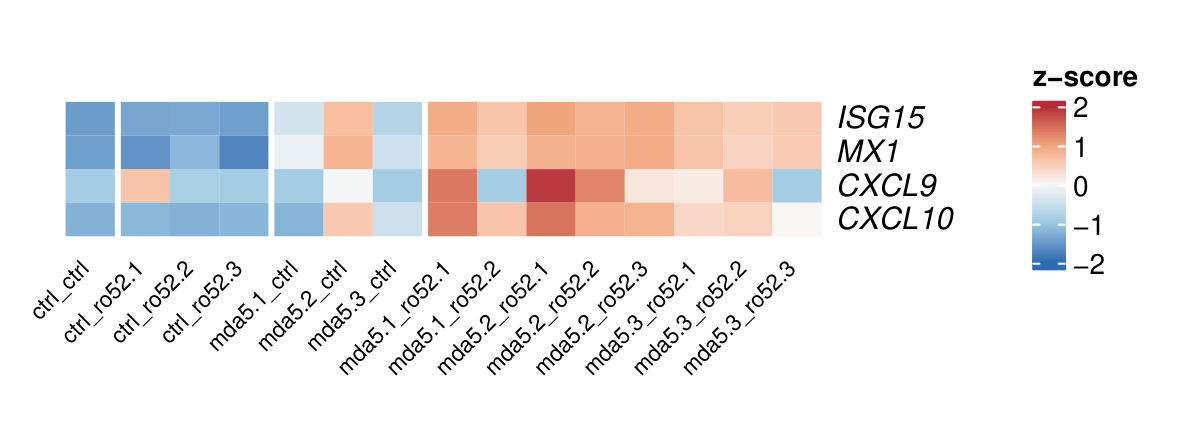

**Supplementary Figure S3. IFN-beta-predominant genes in the MDA5 validation experiment.** The first 50 IFN-beta-predominant genes from the independently defined, P-value-ranked list in Supplementary Table S1 are shown for the validation experiment in Figure 1B.

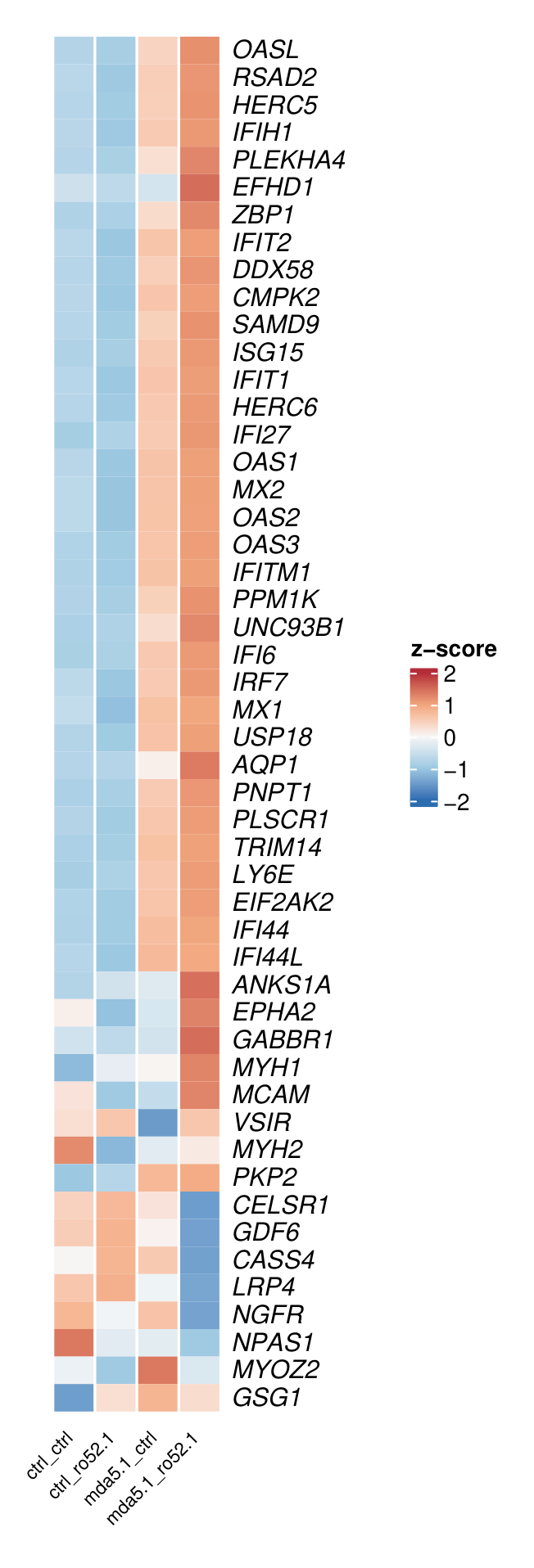

**Supplementary Figure S4. IFN-gamma-predominant genes in the MDA5 validation experiment.** The first 50 IFN-gamma-predominant genes from the independently defined, P-value-ranked list in Supplementary Table S1 are shown for the validation experiment in Figure 1B.

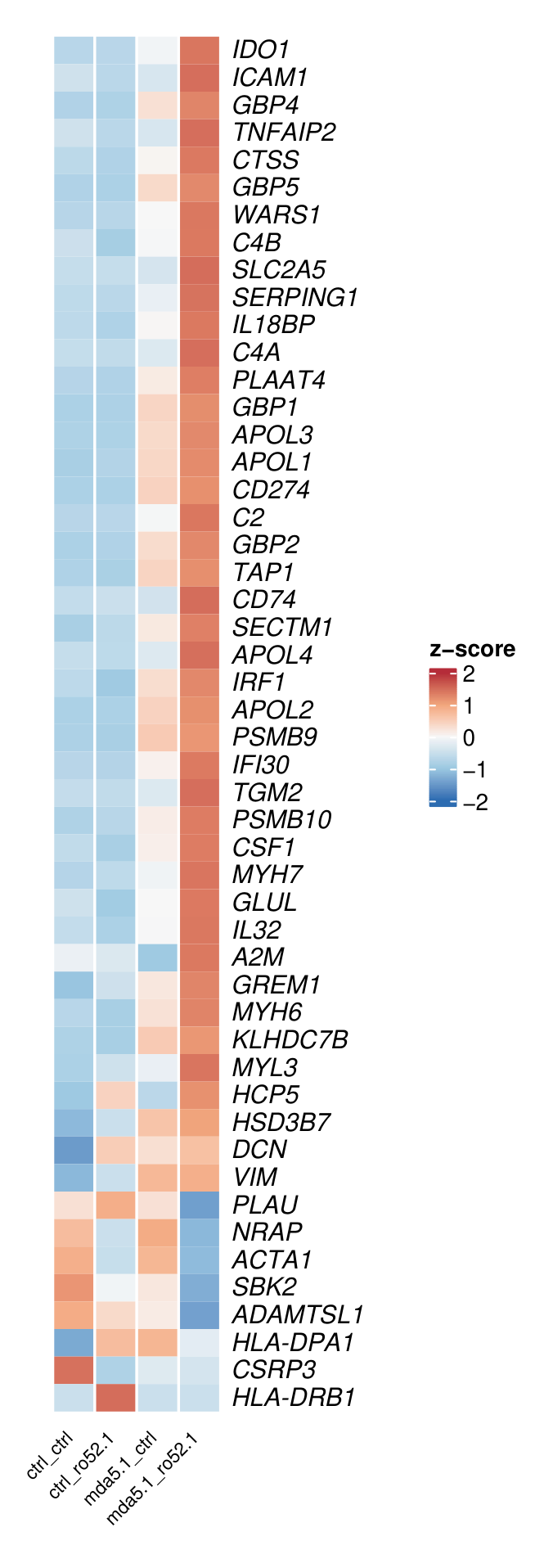

**Supplementary Figure S5. Donor-detail interferon plots for affinity-purified anti-Ro52.** Myoblasts received healthy-control or anti-MDA5-positive IgG with control IgG or anti-Ro52 affinity-purified against full-length Ro52.

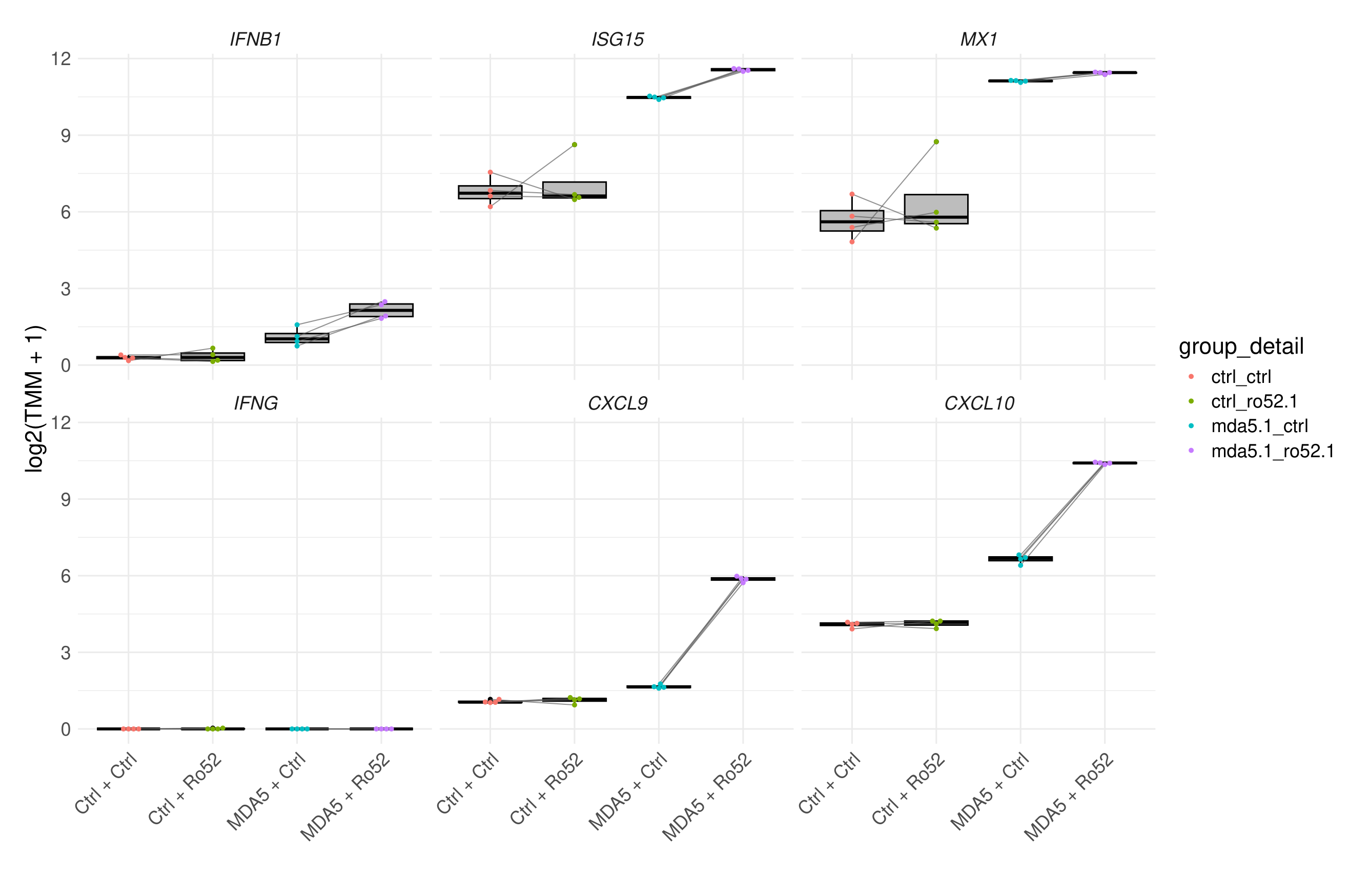

**Supplementary Figure S6. Pathway enrichment after co-electroporation of affinity-purified anti-Ro52 and anti-MDA5.** Anti-MDA5 plus affinity-purified anti-Ro52 was compared with anti-MDA5 plus control IgG. (A) Reactome enrichment: positive normalized enrichment scores favor anti-Ro52; dot size indicates leading-edge gene count and color indicates q-value. (B) Enrichment map: nodes are pathways, sized by leading-edge gene count and colored by -log10(q-value); edges indicate shared genes.

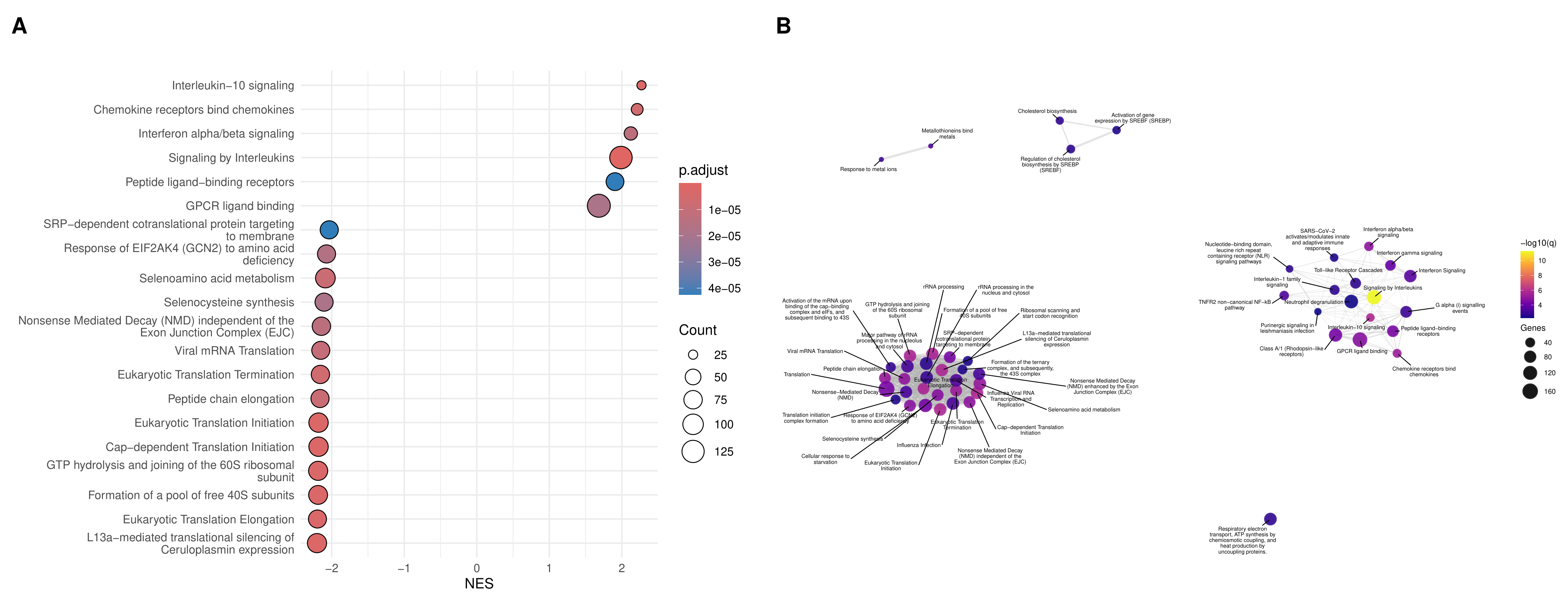

**Supplementary Figure S7. *MX1* qPCR with anti-Ro52 purified against the N-terminal fragment.** Myoblasts were co-electroporated with anti-MDA5-positive IgG and the indicated N-terminal-Ro52-affinity-purified preparation. *MX1* RNA was measured 24 hours later by qPCR and normalized to 18S rRNA.

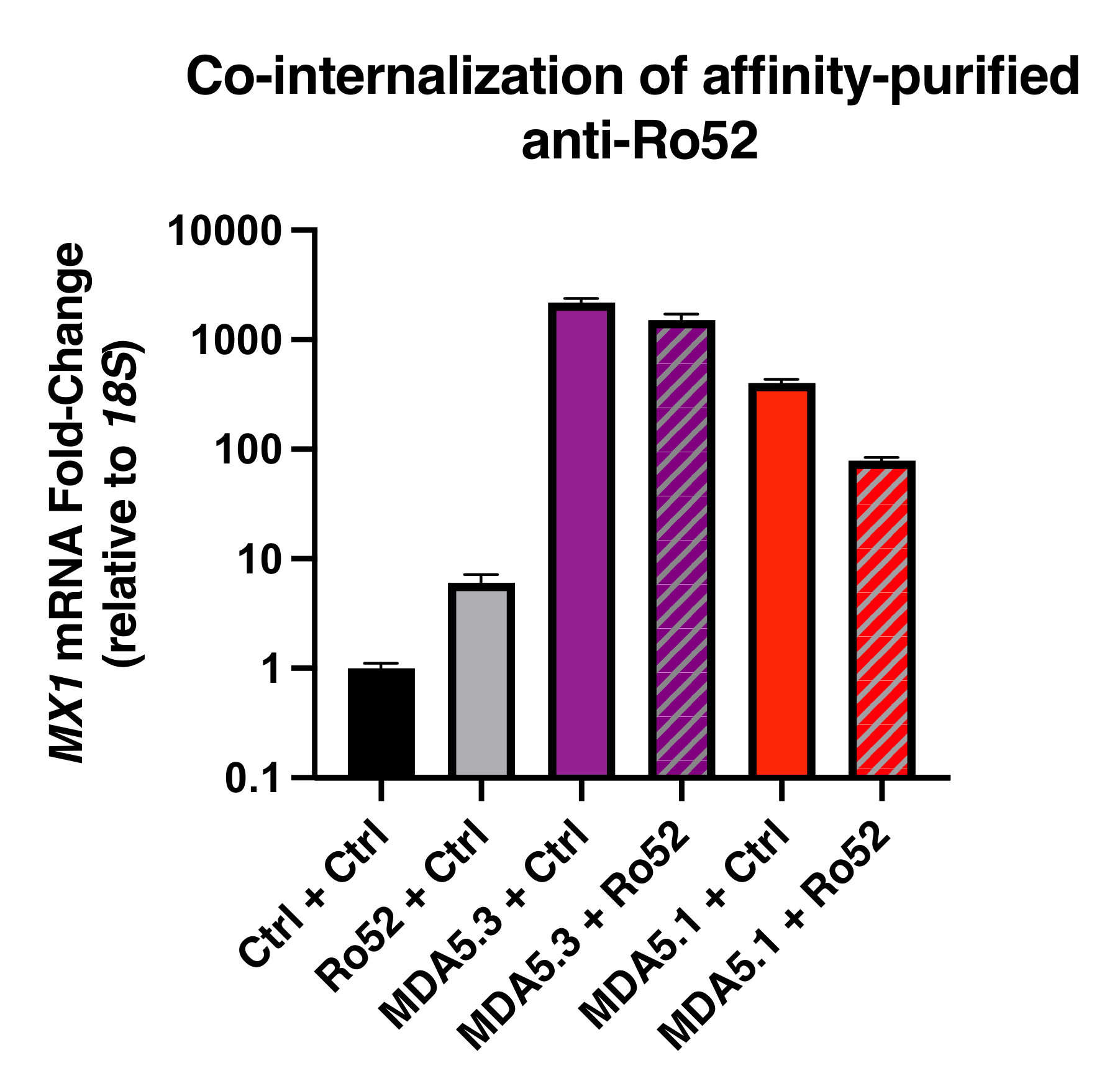

**Supplementary Figure S8. Donor-detail interferon plots for the multi-antibody experiment.** The samples and genes are the same as in Figure 4. Myoblasts received the indicated primary IgG with control IgG or anti-Ro52-positive IgG from patients with myositis, Sjogren disease, or systemic sclerosis.

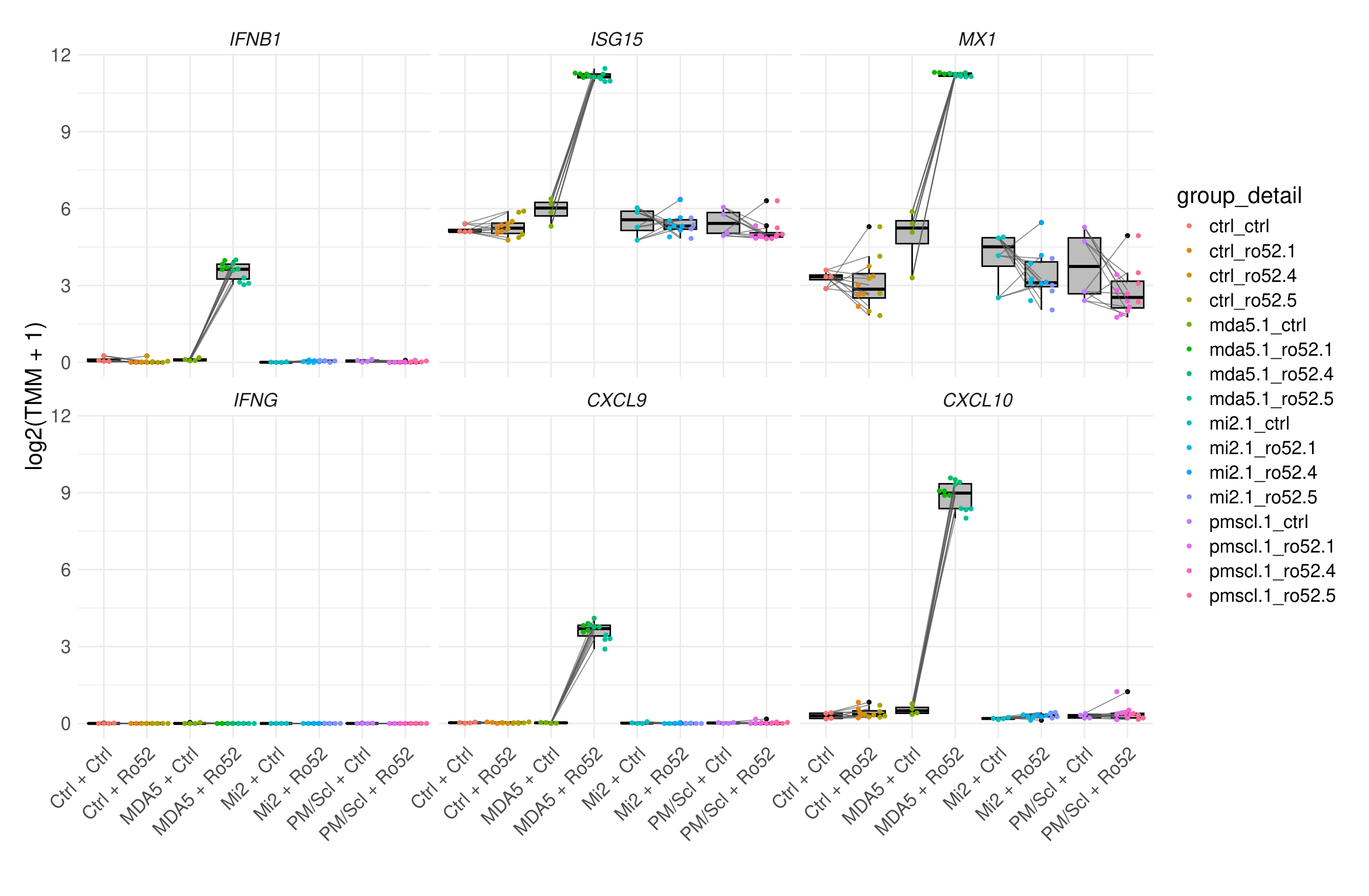

**Supplementary Figure S9. IFN-predominant transcriptional responses in the multi-antibody experiment.** (A) The first 25 IFN-beta-predominant genes. (B) The first 25 IFN-gamma-predominant genes. Gene lists were defined independently and ranked as in Supplementary Table S1.

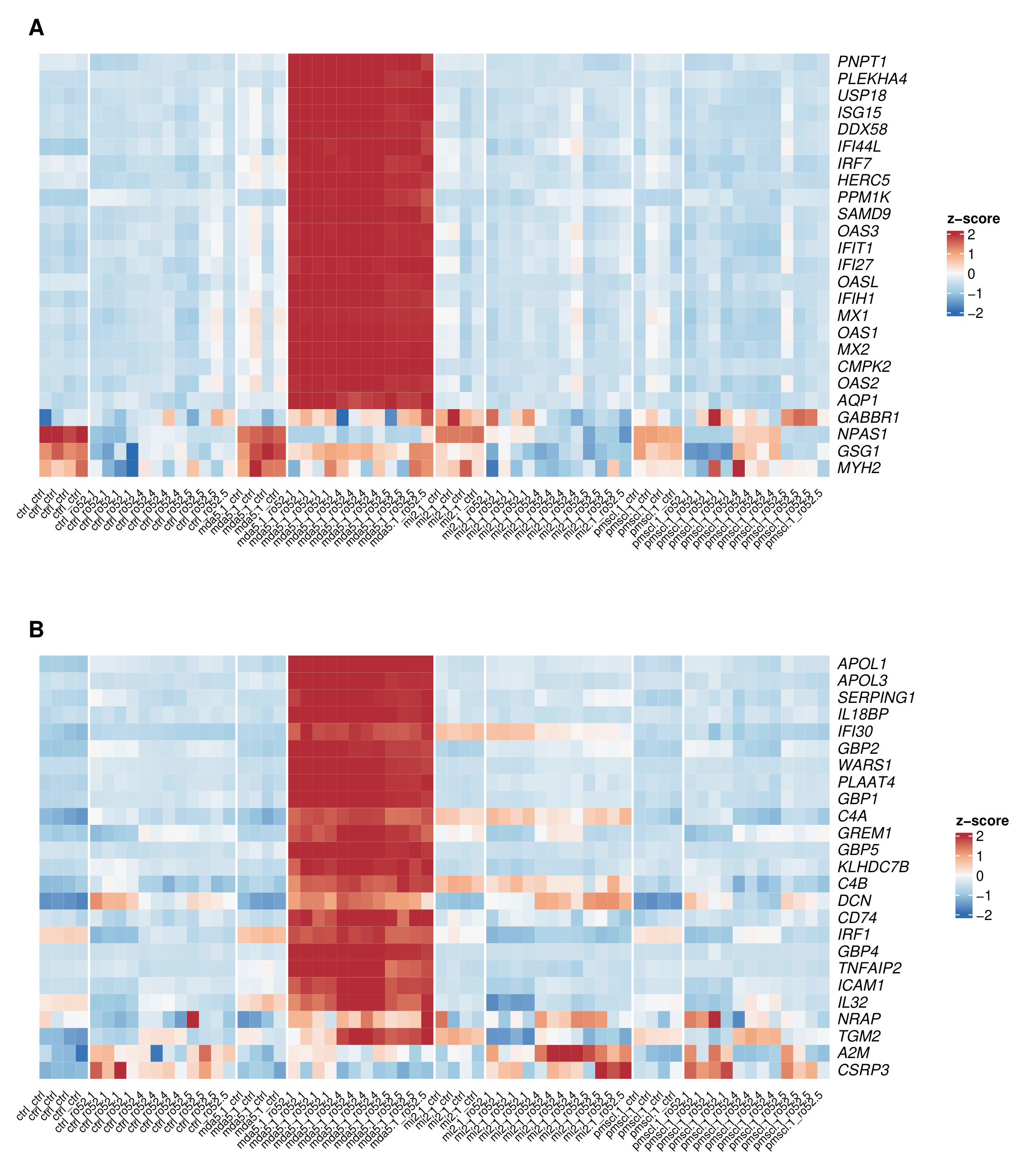

**Supplementary Figure S10. Mi-2- and PM/Scl-associated transcripts in the initial multi-antibody experiment.** *CAMKV*, *EFNA3*, *ENSG00000288751.1*, and *ENSG00000276216.1* expression in human skeletal muscle myoblasts 24 hours after co-electroporation with the indicated IgG combinations.

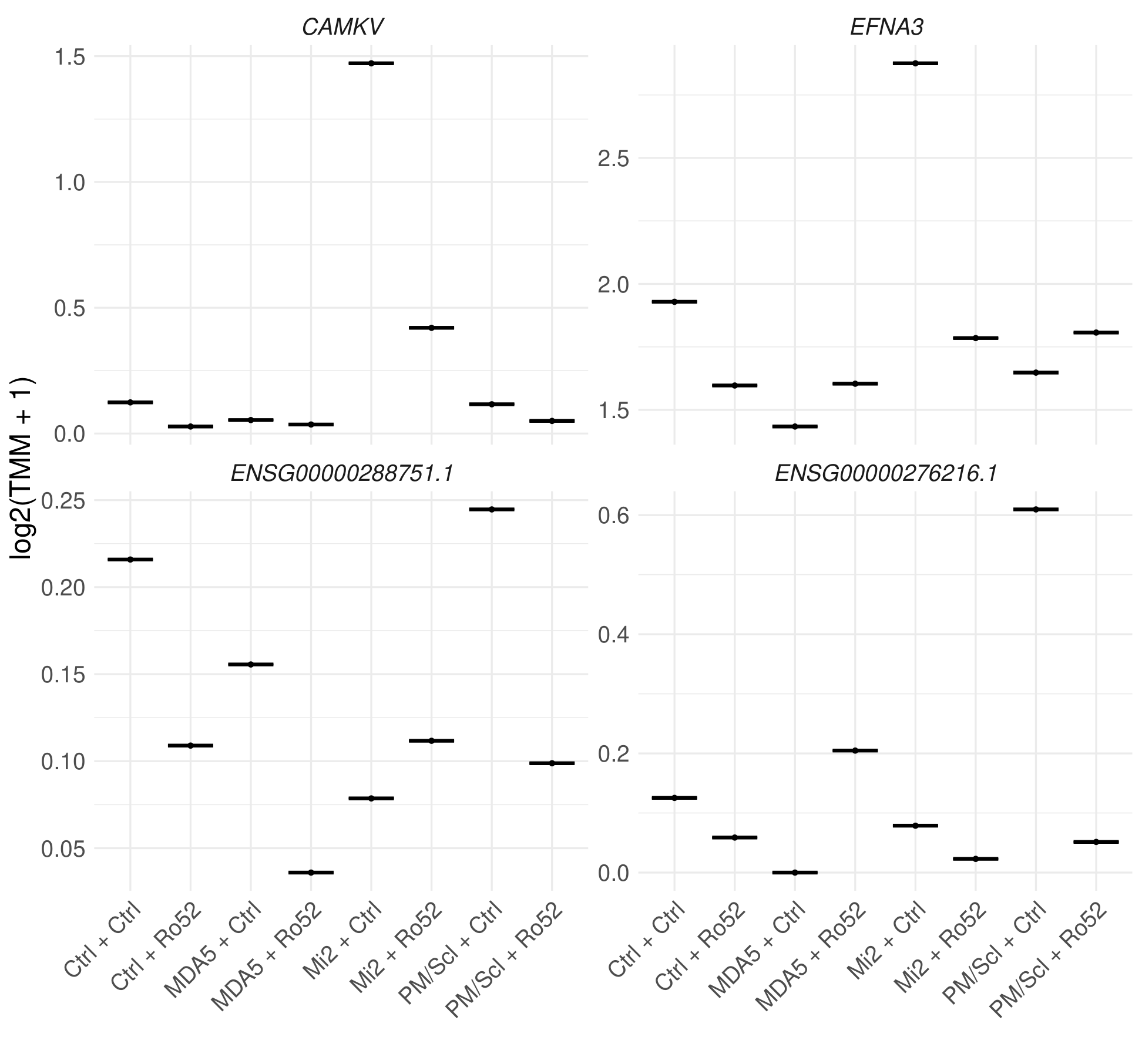

**Supplementary Figure S11. Donor-detail plots for Mi-2- and PM/Scl-associated transcripts.** *CAMKV*, *EFNA3*, *ENSG00000288751.1*, and *ENSG00000276216.1* expression in the samples shown in Figure 7.

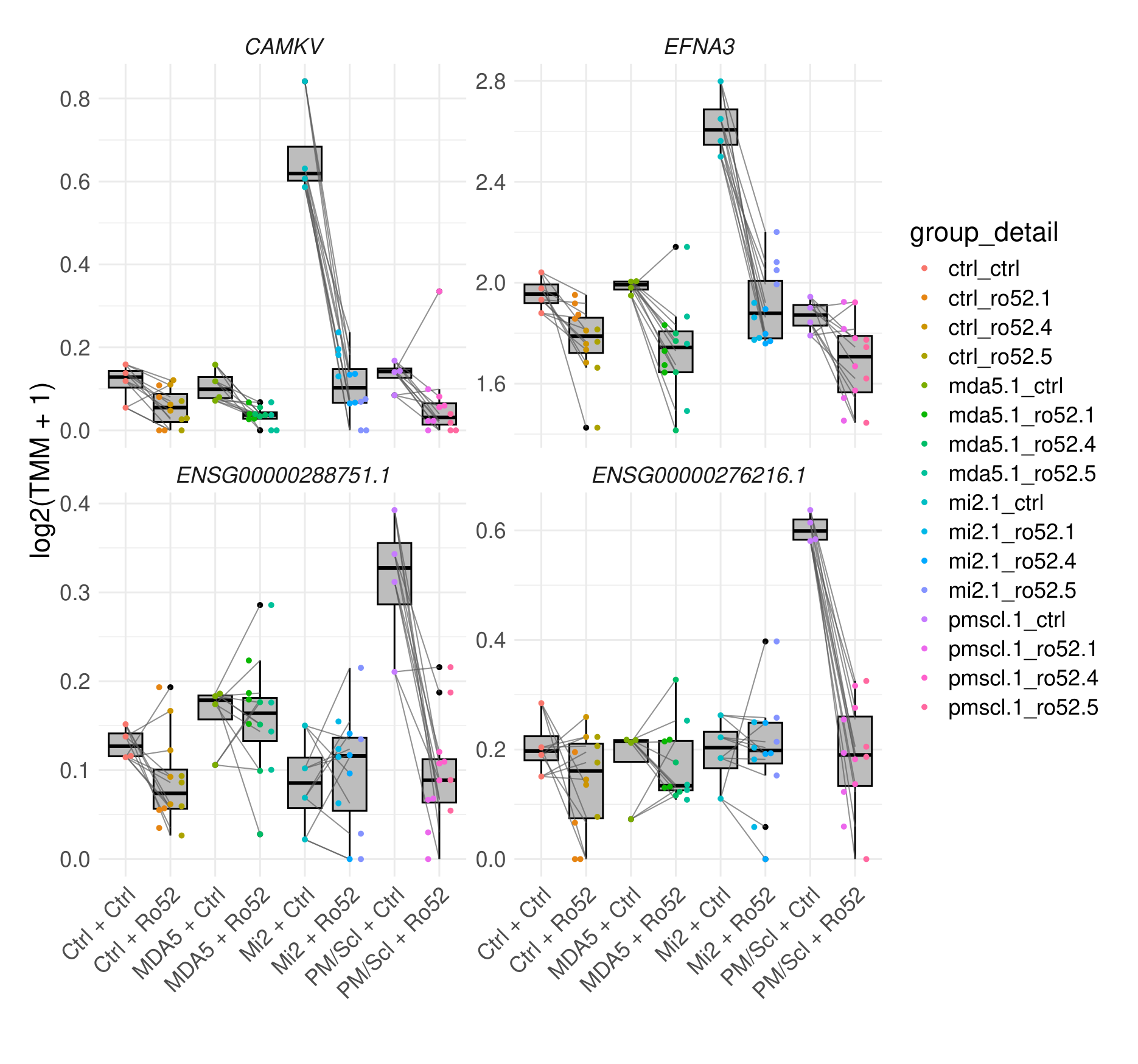
