## Supplementary Tables for "Anti-Ro52 autoantibodies modulate the transcriptomic effects of autoantibodies targeting intracellular autoantigens"

**Supplementary Table S1. Interferon-predominant genes displayed in the figures.** IFN-beta-predominant (A) and IFN-gamma-predominant (B) genes are restricted to those shown in the manuscript. Predominant genes were upregulated relative to both untreated cells and the other IFN, with q<0.05 in both qualifying comparisons. Genes retain ascending raw P-value order from the direct IFN-beta versus IFN-gamma comparison.

**Supplementary Table S1A. IFN-beta-predominant genes**

| **Gene** | **IFN-beta vs. IFN-gamma** | | **IFN-beta vs. untreated** | | **IFN-gamma vs. untreated** | |
| --- | --- | --- | --- | --- | --- | --- |
|  | **log2FC** | **q-value** | **log2FC** | **q-value** | **log2FC** | **q-value** |
| *IFIT1* | 4.342 | 5.65e-13 | 5.616 | 4.69e-13 | 1.285 | 7.74e-07 |
| *MX1* | 3.383 | 8.84e-13 | 6.012 | 4.69e-13 | 2.640 | 3.46e-09 |
| *OAS1* | 3.180 | 9.04e-13 | 5.673 | 4.59e-12 | 2.506 | 2.3e-08 |
| *MX2* | 3.171 | 1.7e-12 | 5.450 | 1.62e-11 | 2.291 | 1.04e-07 |
| *HERC5* | 3.837 | 5.54e-12 | 6.194 | 1.61e-09 | 2.369 | 1.92e-05 |
| *ISG15* | 3.087 | 6.42e-12 | 4.606 | 4.07e-12 | 1.532 | 1.53e-08 |
| *AQP1* | 2.678 | 1.58e-11 | 0.510 | 4.54e-05 | -2.158 | 2.16e-09 |
| *OAS3* | 2.594 | 1.58e-11 | 4.830 | 2.53e-12 | 2.248 | 1.96e-09 |
| *NPAS1* | 2.606 | 2.03e-11 | 1.814 | 1.73e-09 | -0.778 | 6.99e-06 |
| *OASL* | 2.518 | 2.8e-11 | 8.134 | 1.03e-08 | 5.628 | 8.28e-07 |
| *GSG1* | 3.467 | 3.82e-11 | 1.243 | 7.44e-08 | -2.213 | 5.14e-08 |
| *IFI27* | 2.401 | 4.69e-11 | 4.553 | 4.1e-12 | 2.165 | 9.94e-10 |
| *IRF7* | 2.480 | 5.08e-11 | 2.498 | 3.04e-10 | 0.028 | 0.781 |
| *PNPT1* | 2.117 | 5.09e-11 | 2.277 | 3.04e-10 | 0.174 | 0.0395 |
| *IFIH1* | 1.989 | 6.35e-11 | 4.882 | 7.59e-12 | 2.906 | 5.51e-09 |
| *SAMD9* | 2.337 | 6.75e-11 | 3.244 | 4.12e-11 | 0.920 | 3.97e-07 |
| *USP18* | 1.911 | 8.27e-11 | 3.574 | 1.69e-11 | 1.674 | 3.29e-08 |
| *DDX58* | 1.887 | 9.03e-11 | 3.719 | 7.59e-12 | 1.845 | 1.04e-08 |
| *OAS2* | 1.941 | 9.77e-11 | 4.410 | 4.07e-12 | 2.481 | 2.24e-09 |
| *PLEKHA4* | 1.919 | 1.27e-10 | 1.874 | 9.54e-10 | -0.035 | 0.596 |
| *PPM1K* | 1.829 | 1.52e-10 | 1.469 | 1.8e-08 | -0.349 | 0.00083 |
| *IFI44L* | 2.004 | 1.89e-10 | 4.126 | 4.14e-12 | 2.138 | 2.45e-08 |
| *CMPK2* | 1.641 | 9.25e-10 | 8.558 | 1.97e-08 | 6.930 | 4.03e-07 |
| *MYH2* | 1.554 | 9.61e-10 | 0.688 | 3.4e-06 | -0.853 | 1.51e-07 |
| *GABBR1* | 2.036 | 9.65e-10 | 1.246 | 5.4e-07 | -0.775 | 5.34e-05 |
| *IFI44* | 1.595 | 1.09e-09 | 3.057 | 8.66e-11 | 1.475 | 9.91e-08 |
| *IFI6* | 1.690 | 1.17e-09 | 3.693 | 8.35e-12 | 2.015 | 3.59e-10 |
| *TRIM14* | 1.639 | 1.2e-09 | 3.099 | 1.99e-10 | 1.470 | 4.24e-07 |
| *RSAD2* | 1.520 | 1.25e-09 | 10.908 | 1.98e-09 | 9.398 | 1.48e-07 |
| *CELSR1* | 2.598 | 1.36e-09 | 1.195 | 6.99e-07 | -1.390 | 2.37e-06 |
| *NGFR* | 1.430 | 1.37e-09 | 0.261 | 0.00279 | -1.157 | 2.37e-08 |
| *IFIT2* | 1.386 | 1.65e-09 | 5.918 | 2.53e-12 | 4.545 | 3.59e-10 |
| *HERC6* | 1.494 | 1.7e-09 | 3.318 | 1.25e-10 | 1.837 | 6.81e-08 |
| *EFHD1* | 1.562 | 1.71e-09 | 0.424 | 0.0004 | -1.126 | 2.47e-07 |
| *PKP2* | 1.993 | 2.52e-09 | 1.928 | 2.85e-08 | -0.052 | 0.704 |
| *LRP4* | 1.815 | 2.71e-09 | 0.305 | 0.00839 | -1.497 | 4.9e-09 |
| *CASS4* | 1.435 | 2.71e-09 | 0.317 | 0.00171 | -1.105 | 1.84e-07 |
| *MCAM* | 1.347 | 2.88e-09 | 0.642 | 5.89e-06 | -0.694 | 2.6e-06 |
| *EPHA2* | 1.536 | 3.04e-09 | 0.652 | 1.14e-05 | -0.871 | 1.63e-06 |
| *PLSCR1* | 1.304 | 3.27e-09 | 3.045 | 7.02e-11 | 1.753 | 2.52e-08 |
| *ANKS1A* | 1.292 | 3.89e-09 | 0.374 | 0.000236 | -0.906 | 1.01e-07 |
| *GDF6* | 1.378 | 4.04e-09 | 0.409 | 0.000209 | -0.956 | 3.06e-07 |
| *MYH1* | 1.326 | 6.05e-09 | 0.539 | 3.31e-05 | -0.776 | 2.14e-06 |
| *IFITM1* | 1.243 | 8.52e-09 | 5.489 | 3.05e-12 | 4.260 | 1.04e-10 |
| *VSIR* | 1.182 | 8.72e-09 | 0.531 | 1.7e-05 | -0.640 | 2.5e-06 |
| *UNC93B1* | 1.320 | 1.11e-08 | 2.064 | 3.17e-09 | 0.757 | 2.13e-05 |
| *LY6E* | 1.226 | 1.15e-08 | 2.349 | 2.51e-10 | 1.135 | 3.29e-08 |
| *MYOZ2* | 1.106 | 1.76e-08 | 0.458 | 6.3e-05 | -0.635 | 1.61e-06 |
| *EIF2AK2* | 1.286 | 1.8e-08 | 1.612 | 1.29e-08 | 0.339 | 0.000405 |
| *ZBP1* | 3.887 | 1.99e-08 | 9.056 | 1.28e-05 | 5.182 | 0.000604 |

**Supplementary Table S1B. IFN-gamma-predominant genes**

| **Gene** | **IFN-beta vs. IFN-gamma** | | **IFN-beta vs. untreated** | | **IFN-gamma vs. untreated** | |
| --- | --- | --- | --- | --- | --- | --- |
|  | **log2FC** | **q-value** | **log2FC** | **q-value** | **log2FC** | **q-value** |
| *CD74* | -9.172 | 5.65e-13 | 2.855 | 3.47e-05 | 12.041 | 1.46e-09 |
| *WARS1* | -3.520 | 5.65e-13 | 0.894 | 1.78e-07 | 4.427 | 9.65e-12 |
| *ICAM1* | -4.447 | 8.2e-13 | -1.139 | 2.91e-07 | 3.321 | 6.26e-11 |
| *TNFAIP2* | -3.374 | 8.84e-13 | -0.427 | 0.00024 | 2.959 | 1.25e-10 |
| *GBP5* | -8.128 | 8.84e-13 | 4.878 | 3.13e-05 | 13.019 | 7.62e-08 |
| *IRF1* | -3.691 | 8.84e-13 | 1.402 | 1.09e-07 | 5.105 | 3.11e-11 |
| *PLAAT4* | -3.705 | 9.04e-13 | 2.645 | 2.29e-08 | 6.363 | 2.27e-10 |
| *GBP1* | -3.037 | 9.44e-13 | 2.528 | 1.45e-10 | 5.578 | 1.36e-11 |
| *GBP4* | -4.368 | 1.04e-12 | 6.612 | 1.49e-06 | 10.993 | 3.2e-08 |
| *GBP2* | -3.138 | 1.1e-12 | 1.512 | 1.31e-08 | 4.661 | 1.81e-11 |
| *TGM2* | -2.680 | 6.42e-12 | -0.507 | 0.000412 | 2.187 | 1.29e-08 |
| *NRAP* | -3.306 | 6.99e-12 | 0.556 | 0.00133 | 3.873 | 6.99e-10 |
| *A2M* | -3.046 | 1.39e-11 | -2.557 | 7.98e-10 | 0.500 | 0.00071 |
| *APOL1* | -2.298 | 1.58e-11 | 3.539 | 1.74e-11 | 5.850 | 1.36e-11 |
| *IL32* | -2.552 | 1.58e-11 | -0.019 | 0.838 | 2.545 | 2.4e-10 |
| *C4A* | -3.089 | 1.7e-11 | 1.933 | 1.53e-08 | 5.034 | 7.99e-10 |
| *IL18BP* | -3.704 | 1.72e-11 | 0.724 | 0.00048 | 4.441 | 4.83e-10 |
| *SERPING1* | -2.281 | 1.76e-11 | 2.346 | 3.75e-10 | 4.639 | 2.94e-11 |
| *CSRP3* | -2.718 | 2.03e-11 | -0.005 | 0.964 | 2.724 | 2.7e-10 |
| *C4B* | -3.960 | 2.13e-11 | 2.566 | 3.96e-07 | 6.538 | 1.07e-08 |
| *DCN* | -2.704 | 2.6e-11 | 0.023 | 0.801 | 2.741 | 9.43e-10 |
| *APOL3* | -2.639 | 2.72e-11 | 3.165 | 1.03e-08 | 5.816 | 4.83e-10 |
| *KLHDC7B* | -3.198 | 3.7e-11 | 3.168 | 2.24e-08 | 6.379 | 4.91e-09 |
| *IFI30* | -2.825 | 6.75e-11 | 3.375 | 5.99e-08 | 6.212 | 9.94e-10 |
| *GREM1* | -2.059 | 6.75e-11 | -1.088 | 3.54e-07 | 0.984 | 6.35e-07 |
| *IDO1* | -9.975 | 7.05e-11 | 3.023 | 0.00127 | 13.013 | 3.85e-07 |
| *PSMB10* | -2.668 | 7.96e-11 | 0.920 | 1.51e-05 | 3.600 | 4.68e-10 |
| *HLA-DPA1* | -7.516 | 8.06e-11 | 0.986 | 0.0353 | 8.517 | 6.28e-09 |
| *MYH6* | -1.962 | 9.77e-11 | 0.606 | 1.04e-05 | 2.580 | 2.7e-10 |
| *SECTM1* | -2.637 | 9.77e-11 | 3.413 | 6.93e-08 | 6.064 | 1.86e-08 |
| *APOL4* | -2.320 | 1.12e-10 | -0.303 | 0.0138 | 2.029 | 7.4e-09 |
| *C2* | -1.987 | 1.19e-10 | 2.477 | 2.34e-09 | 4.477 | 2.57e-10 |
| *CTSS* | -4.362 | 1.19e-10 | 2.818 | 5.97e-06 | 7.193 | 5.99e-08 |
| *APOL2* | -1.906 | 1.44e-10 | 1.816 | 1.88e-09 | 3.735 | 2.82e-11 |
| *TAP1* | -1.817 | 1.44e-10 | 3.401 | 7.59e-12 | 5.230 | 1.36e-11 |
| *PSMB9* | -2.016 | 1.63e-10 | 3.984 | 2.85e-09 | 6.014 | 7.3e-10 |
| *CSF1* | -1.818 | 1.79e-10 | 0.984 | 2.22e-07 | 2.814 | 1.84e-10 |
| *VIM* | -1.702 | 1.79e-10 | -1.320 | 4.94e-09 | 0.395 | 1.62e-05 |
| *PLAU* | -2.070 | 1.95e-10 | -0.808 | 2.36e-06 | 1.274 | 7.58e-08 |
| *GLUL* | -1.726 | 2.36e-10 | 0.205 | 0.0136 | 1.944 | 9.43e-10 |
| *HLA-DRB1* | -9.707 | 2.55e-10 | 3.931 | 0.0121 | 13.653 | 9.66e-07 |
| *HCP5* | -2.018 | 2.58e-10 | 7.246 | 2.77e-07 | 9.276 | 9.73e-08 |
| *CD274* | -2.542 | 2.62e-10 | 1.146 | 1.5e-05 | 3.701 | 2.33e-09 |
| *HSD3B7* | -2.420 | 2.71e-10 | 0.010 | 0.946 | 2.442 | 2.41e-09 |
| *MYL3* | -2.032 | 2.8e-10 | -0.889 | 3.81e-05 | 1.153 | 1.72e-06 |
| *MYH7* | -1.973 | 4.86e-10 | 0.391 | 0.000123 | 2.377 | 2.45e-09 |
| *ADAMTSL1* | -1.905 | 6.26e-10 | -1.014 | 1.47e-06 | 0.905 | 8.74e-07 |
| *SBK2* | -2.157 | 6.8e-10 | 0.567 | 0.00053 | 2.735 | 4.9e-09 |
| *SLC2A5* | -2.101 | 7.13e-10 | 0.951 | 7.9e-05 | 3.063 | 1.01e-08 |
| *ACTA1* | -1.649 | 7.38e-10 | -0.311 | 0.0112 | 1.350 | 2.34e-07 |
| *CXCL10* | -4.468 | 3.82e-09 | 5.893 | 0.000206 | 10.376 | 1.14e-05 |
| *CXCL9* | -11.986 | 1.38e-08 | 2.796 | 0.0983 | 14.804 | 2.21e-06 |

**Supplementary Table S2. Effects of affinity-purified anti-Ro52 on anti-MDA5-associated gene expression.** For each primary autoantibody, IgG co-electroporated with anti-Ro52 was compared with the same primary IgG plus control IgG (4 versus 4 RNA-seq libraries per comparison). The table includes the 108 distinct genes displayed in the main and supplementary expression figures. Blank cells indicate insufficient gene expression for inclusion in differential expression analysis. Anti-Ro52 was affinity-purified using full-length Ro52; the MDA5 and Ro52 preparations each originate from one IgG donor.

| **Gene** | **Anti-MDA5 + Ro52 vs. Anti-MDA5 + control** | |
| --- | --- | --- |
|  | **log2FC** | **q-value** |
| *IFNB1* | 1.628 | 0.0115 |
| *ISG15* | 1.065 | 9.66e-07 |
| *MX1* | 0.301 | 2.4e-05 |
| *IFNG* |  |  |
| *CXCL9* | 4.697 | 5.56e-07 |
| *CXCL10* | 3.752 | 8.88e-08 |
| *CAMKV* | 0.205 | 1 |
| *EFNA3* | 0.281 | 0.143 |
| *ENSG00000288751.1* | -1.340 | 0.26 |
| *ENSG00000276216.1* | 1.093 | 0.585 |
| *IFIH1* | 0.628 | 4.44e-06 |
| *IFIT1* | 0.986 | 2.72e-07 |
| *OAS1* | 0.730 | 1.41e-06 |
| *MX2* | 0.485 | 0.000287 |
| *HERC5* | 0.460 | 0.000607 |
| *AQP1* | -0.421 | 0.00071 |
| *OAS3* | 0.512 | 3.81e-05 |
| *NPAS1* | -0.378 | 0.112 |
| *OASL* | 1.736 | 2.04e-06 |
| *GSG1* | -0.146 | 0.437 |
| *IFI27* | 0.871 | 1.87e-05 |
| *IRF7* | 1.046 | 1.95e-07 |
| *PNPT1* | 0.106 | 0.000597 |
| *SAMD9* | 0.652 | 8.57e-06 |
| *USP18* | 0.569 | 7.41e-06 |
| *DDX58* | 0.738 | 1.53e-06 |
| *OAS2* | 0.378 | 0.00044 |
| *PLEKHA4* | 0.619 | 3.66e-06 |
| *PPM1K* | 0.152 | 0.124 |
| *IFI44L* | -0.131 | 0.00256 |
| *CMPK2* | 0.874 | 1.64e-06 |
| *MYH2* | 0.084 | 0.611 |
| *GABBR1* | -0.202 | 0.0313 |
| *IFI44* | 0.010 | 0.935 |
| *IFI6* | 0.907 | 4.96e-06 |
| *TRIM14* | 0.001 | 1 |
| *RSAD2* | 1.734 | 4.27e-07 |
| *CELSR1* | -0.695 | 0.056 |
| *NGFR* | 0.042 | 0.682 |
| *IFIT2* | 0.823 | 1.21e-05 |
| *HERC6* | 0.649 | 4.87e-06 |
| *EFHD1* | 1.773 | 0.000113 |
| *PKP2* | -0.584 | 0.397 |
| *LRP4* | -0.661 | 6.05e-06 |
| *CASS4* | -0.043 | 0.968 |
| *MCAM* | 0.718 | 3.02e-06 |
| *EPHA2* | 0.107 | 0.287 |
| *PLSCR1* | 0.400 | 1.54e-05 |
| *ANKS1A* | -0.198 | 0.00543 |
| *GDF6* | -1.074 | 0.0035 |
| *MYH1* | 0.264 | 0.0642 |
| *IFITM1* | 0.795 | 1.16e-05 |
| *VSIR* | 0.501 | 4.19e-06 |
| *UNC93B1* | 0.848 | 1.06e-06 |
| *LY6E* | 0.603 | 1.59e-05 |
| *MYOZ2* | 0.522 | 0.115 |
| *EIF2AK2* | -0.226 | 0.000436 |
| *ZBP1* | 1.048 | 0.000343 |
| *CD74* | 0.770 | 4.33e-05 |
| *WARS1* | 1.311 | 1.01e-05 |
| *ICAM1* | 2.737 | 4.91e-07 |
| *TNFAIP2* | 2.354 | 1.49e-06 |
| *GBP5* | 1.620 | 2.68e-06 |
| *IRF1* | 0.915 | 4.51e-05 |
| *PLAAT4* | 1.251 | 4.16e-06 |
| *GBP1* | 1.116 | 1.34e-06 |
| *GBP4* | 1.444 | 2.59e-06 |
| *GBP2* | 0.508 | 0.000186 |
| *TGM2* | 0.264 | 8.75e-05 |
| *NRAP* | 0.280 | 0.073 |
| *A2M* | -0.178 | 0.176 |
| *APOL1* | 1.295 | 8.39e-07 |
| *IL32* | 1.766 | 5.37e-08 |
| *C4A* | 0.505 | 0.00284 |
| *IL18BP* | 1.321 | 9.66e-07 |
| *SERPING1* | 1.258 | 1.32e-05 |
| *CSRP3* | 0.024 | 1 |
| *C4B* | 0.449 | 0.0703 |
| *DCN* | 0.302 | 6.63e-05 |
| *APOL3* | 1.039 | 3.06e-07 |
| *KLHDC7B* | 1.102 | 3.59e-05 |
| *IFI30* | 1.230 | 4.7e-07 |
| *GREM1* | 0.582 | 6.24e-06 |
| *IDO1* | 2.969 | 1.16e-06 |
| *PSMB10* | 0.898 | 9.43e-06 |
| *HLA-DPA1* | 0.593 | 0.000222 |
| *MYH6* | -0.008 | 1 |
| *SECTM1* | 1.063 | 6.25e-07 |
| *APOL4* | 0.557 | 0.0466 |
| *C2* | 1.236 | 6.21e-06 |
| *CTSS* | 2.957 | 4.96e-07 |
| *APOL2* | 0.874 | 2.29e-05 |
| *TAP1* | 0.709 | 4.63e-07 |
| *PSMB9* | 0.664 | 2.05e-05 |
| *CSF1* | 0.653 | 4.04e-07 |
| *VIM* | -0.400 | 9.83e-07 |
| *PLAU* | -0.419 | 0.00011 |
| *GLUL* | 0.167 | 0.00397 |
| *HLA-DRB1* | 0.178 | 0.347 |
| *HCP5* | 1.481 | 0.000176 |
| *CD274* | 0.799 | 0.000701 |
| *HSD3B7* | 0.595 | 3.22e-05 |
| *MYL3* | -0.004 | 1 |
| *MYH7* | 0.201 | 0.325 |
| *ADAMTSL1* | -0.345 | 4.38e-05 |
| *SBK2* | -2.399 | 7.64e-06 |
| *SLC2A5* | 2.239 | 1.67e-05 |
| *ACTA1* | 0.130 | 0.183 |

**Supplementary Table S3. Effects of anti-Ro52 IgG from multiple autoimmune diseases on autoantibody-associated gene expression.** For each primary autoantibody, IgG co-electroporated with anti-Ro52 IgG was compared with the same primary IgG co-electroporated with control IgG (12 versus 4 RNA-seq libraries, respectively, per comparison). The table includes the 108 distinct genes shown in the manuscript. Blank cells indicate insufficient gene expression for inclusion in differential expression analysis. Anti-Ro52 IgG was obtained from patients with myositis, Sjögren disease, or systemic sclerosis.

| **Gene** | **Anti-MDA5 + Ro52 vs. Anti-MDA5 + control** | | **Anti-Mi-2 + Ro52 vs. Anti-Mi-2 + control** | | **Anti-PM/Scl + Ro52 vs. Anti-PM/Scl + control** | |
| --- | --- | --- | --- | --- | --- | --- |
|  | **log2FC** | **q-value** | **log2FC** | **q-value** | **log2FC** | **q-value** |
| *IFNB1* | 7.230 | 2.29e-09 | 2.134 | 0.0092 | -1.042 | 0.144 |
| *ISG15* | 5.295 | 2.14e-16 | -0.108 | 0.686 | -0.329 | 0.235 |
| *MX1* | 6.430 | 3.41e-14 | -0.781 | 0.139 | -1.079 | 0.134 |
| *IFNG* |  |  |  |  |  |  |
| *CXCL9* | 9.712 | 1.57e-08 | 0.191 | 0.832 | 0.929 | 0.366 |
| *CXCL10* | 10.189 | 1.4e-07 | 0.712 | 0.0252 | 0.370 | 0.494 |
| *CAMKV* | -1.064 | 0.00259 | -2.995 | 6.37e-07 | -1.539 | 0.0358 |
| *EFNA3* | -0.322 | 0.00526 | -0.946 | 4.48e-08 | -0.253 | 0.0382 |
| *ENSG00000288751.1* | -0.039 | 0.924 | 0.454 | 0.481 | -1.954 | 0.000688 |
| *ENSG00000276216.1* | 0.097 | 0.795 | -0.033 | 0.955 | -2.021 | 0.000157 |
| *IFIH1* | 6.346 | 2e-14 | 0.050 | 0.887 | -0.334 | 0.458 |
| *IFIT1* | 6.681 | 6.8e-15 | 0.601 | 0.0504 | 0.005 | 0.994 |
| *OAS1* | 6.927 | 4.54e-14 | -0.161 | 0.85 | -0.710 | 0.524 |
| *MX2* | 6.554 | 5.3e-14 | -0.490 | 0.472 | -0.982 | 0.268 |
| *HERC5* | 6.592 | 6.86e-16 | -0.638 | 0.00548 | -1.128 | 0.000362 |
| *AQP1* | 1.574 | 1.46e-13 | -0.103 | 0.2 | -0.038 | 0.701 |
| *OAS3* | 5.691 | 2.14e-15 | -0.333 | 0.447 | -0.381 | 0.479 |
| *NPAS1* | -0.947 | 4.03e-11 | -0.852 | 9.65e-08 | -0.598 | 0.000231 |
| *OASL* | 7.887 | 1.24e-14 | -0.300 | 0.11 | -0.829 | 0.00766 |
| *GSG1* | -0.247 | 0.000695 | -0.323 | 0.000248 | -0.368 | 0.0046 |
| *IFI27* | 5.973 | 9.24e-15 | -0.229 | 0.644 | -0.002 | 0.998 |
| *IRF7* | 3.478 | 4.59e-16 | -0.478 | 0.00415 | -0.756 | 0.000473 |
| *PNPT1* | 3.352 | 4.66e-19 | -0.358 | 5.23e-05 | -0.310 | 0.00237 |
| *SAMD9* | 4.237 | 1.49e-15 | -0.045 | 0.796 | -0.294 | 0.191 |
| *USP18* | 5.432 | 2.13e-16 | 0.142 | 0.52 | -0.117 | 0.73 |
| *DDX58* | 5.219 | 2.73e-16 | 0.149 | 0.324 | -0.236 | 0.198 |
| *OAS2* | 6.521 | 9.33e-14 | -0.230 | 0.726 | -0.754 | 0.41 |
| *PLEKHA4* | 3.950 | 8.63e-19 | 0.046 | 0.346 | -0.003 | 0.978 |
| *PPM1K* | 3.212 | 7.63e-16 | 0.330 | 0.0252 | 0.334 | 0.00462 |
| *IFI44L* | 3.887 | 3.88e-16 | -0.215 | 0.352 | 0.065 | 0.797 |
| *CMPK2* | 7.499 | 8.41e-14 | 0.055 | 0.865 | -0.704 | 0.159 |
| *MYH2* | -0.367 | 0.00555 | -0.468 | 0.00138 | -0.028 | 0.872 |
| *GABBR1* | 0.136 | 0.0527 | -0.194 | 0.0204 | 0.118 | 0.125 |
| *IFI44* | 3.848 | 7.63e-16 | 0.063 | 0.737 | -0.021 | 0.934 |
| *IFI6* | 5.241 | 2.04e-16 | 0.094 | 0.788 | -0.133 | 0.766 |
| *TRIM14* | 4.139 | 1.17e-18 | -0.308 | 0.0242 | -0.401 | 0.041 |
| *RSAD2* | 8.858 | 1.92e-14 | 0.229 | 0.698 | -0.509 | 0.454 |
| *CELSR1* | -0.982 | 1.16e-05 | 0.154 | 0.25 | -0.157 | 0.204 |
| *NGFR* | -0.016 | 0.744 | -0.392 | 9.45e-10 | -0.356 | 3.14e-05 |
| *IFIT2* | 7.621 | 3.42e-15 | 0.418 | 0.00101 | -0.255 | 0.392 |
| *HERC6* | 5.386 | 8.75e-16 | -0.090 | 0.787 | -0.208 | 0.621 |
| *EFHD1* | 2.038 | 7.43e-14 | -0.194 | 0.0438 | -0.124 | 0.417 |
| *PKP2* | 0.241 | 0.445 | -0.812 | 0.00451 | -0.526 | 0.0981 |
| *LRP4* | -0.097 | 0.149 | 0.164 | 0.0791 | 0.150 | 0.0847 |
| *CASS4* | -0.261 | 0.00231 | -0.444 | 0.00161 | -0.071 | 0.484 |
| *MCAM* | -0.083 | 0.0369 | -0.580 | 1.21e-09 | -0.327 | 0.000121 |
| *EPHA2* | -0.905 | 4.12e-11 | -1.053 | 6.01e-13 | -0.915 | 4.85e-05 |
| *PLSCR1* | 3.462 | 9.19e-18 | 0.176 | 0.129 | -0.137 | 0.265 |
| *ANKS1A* | 0.479 | 3.46e-10 | 0.107 | 0.013 | 0.133 | 0.0642 |
| *GDF6* | -1.254 | 3.26e-14 | -0.976 | 6.05e-10 | -0.587 | 0.00271 |
| *MYH1* | 0.592 | 0.00999 | -0.265 | 0.223 | 0.167 | 0.445 |
| *IFITM1* | 6.354 | 1e-14 | 0.143 | 0.737 | 0.109 | 0.862 |
| *VSIR* | -0.139 | 0.0124 | -0.401 | 8.22e-08 | -0.338 | 0.00123 |
| *UNC93B1* | 3.403 | 8.63e-19 | -0.031 | 0.732 | 0.173 | 0.023 |
| *LY6E* | 2.498 | 2.13e-17 | 0.054 | 0.404 | 0.025 | 0.804 |
| *MYOZ2* | -0.849 | 8.33e-07 | -0.642 | 4.58e-05 | -0.399 | 0.0185 |
| *EIF2AK2* | 2.789 | 9.59e-16 | 0.059 | 0.614 | 0.034 | 0.83 |
| *ZBP1* | 6.898 | 1.14e-15 | -1.187 | 0.0252 | -0.616 | 0.464 |
| *CD74* | 2.516 | 2.03e-12 | -0.322 | 0.193 | 0.358 | 0.104 |
| *WARS1* | 3.892 | 1.11e-16 | 0.146 | 0.0286 | 0.179 | 0.000362 |
| *ICAM1* | 1.827 | 1.64e-10 | -0.021 | 0.812 | -0.031 | 0.751 |
| *TNFAIP2* | 1.856 | 9.79e-11 | -0.043 | 0.489 | -0.078 | 0.0478 |
| *GBP5* | 6.838 | 5.31e-14 | -0.325 | 0.304 | -1.164 | 0.0114 |
| *IRF1* | 1.032 | 2.63e-12 | -0.952 | 5.83e-11 | -0.800 | 0.000331 |
| *PLAAT4* | 5.641 | 1.12e-16 | 0.559 | 9.84e-05 | 0.183 | 0.213 |
| *GBP1* | 5.730 | 1.94e-16 | 0.321 | 0.00358 | 0.346 | 0.0453 |
| *GBP4* | 8.871 | 4.27e-12 | 0.613 | 0.151 | 0.249 | 0.716 |
| *GBP2* | 4.544 | 3.66e-17 | 0.969 | 1.23e-07 | 0.683 | 0.0166 |
| *TGM2* | 0.401 | 0.000213 | -0.385 | 1.73e-05 | -0.072 | 0.364 |
| *NRAP* | 2.538 | 3.49e-05 | 0.681 | 0.155 | 0.546 | 0.317 |
| *A2M* | 0.282 | 0.00466 | 0.577 | 0.00128 | 0.364 | 0.0364 |
| *APOL1* | 4.865 | 8.9e-21 | 0.305 | 0.000743 | 0.407 | 0.00549 |
| *IL32* | 0.541 | 1.19e-07 | -0.181 | 0.0262 | -0.154 | 0.0468 |
| *C4A* | 3.176 | 4.73e-16 | -0.056 | 0.716 | 0.218 | 0.27 |
| *IL18BP* | 3.964 | 1.04e-18 | -0.167 | 0.115 | 0.086 | 0.488 |
| *SERPING1* | 2.214 | 8.63e-19 | 0.068 | 0.463 | 0.263 | 0.00189 |
| *CSRP3* | 0.707 | 0.0542 | 1.116 | 0.00978 | 0.984 | 0.0199 |
| *C4B* | 3.444 | 4.05e-13 | -0.521 | 0.0121 | 0.073 | 0.82 |
| *DCN* | 0.758 | 6.74e-13 | 0.484 | 1.51e-06 | 0.447 | 5.12e-05 |
| *APOL3* | 3.918 | 5.7e-19 | 0.000 | 0.998 | -0.131 | 0.165 |
| *KLHDC7B* | 4.719 | 4.05e-13 | -0.456 | 0.278 | 0.234 | 0.424 |
| *IFI30* | 1.687 | 2.64e-17 | -0.210 | 0.0236 | -0.065 | 0.272 |
| *GREM1* | 1.561 | 1.07e-14 | -0.238 | 0.0356 | 0.111 | 0.349 |
| *IDO1* | 6.984 | 7.2e-12 | 1.191 | 1.96e-07 | 0.479 | 0.00588 |
| *PSMB10* | 2.372 | 3.68e-17 | 0.114 | 0.0285 | 0.089 | 0.185 |
| *HLA-DPA1* | 0.838 | 4.97e-10 | 0.277 | 0.0253 | 0.236 | 0.133 |
| *MYH6* | 0.859 | 0.000201 | 0.261 | 0.383 | -0.176 | 0.631 |
| *SECTM1* | 5.269 | 2.36e-20 | 0.666 | 0.00016 | 0.815 | 0.00294 |
| *APOL4* | 2.590 | 2.62e-12 | 0.377 | 0.0203 | 0.270 | 0.231 |
| *C2* | 3.441 | 6.02e-17 | 0.900 | 6.77e-06 | 0.787 | 0.0258 |
| *CTSS* | 4.785 | 6.03e-13 | -0.173 | 0.278 | 0.081 | 0.585 |
| *APOL2* | 3.244 | 6.7e-18 | 0.322 | 0.000137 | 0.256 | 0.00135 |
| *TAP1* | 3.750 | 1.71e-20 | 0.005 | 0.941 | -0.205 | 0.00123 |
| *PSMB9* | 5.581 | 6.45e-19 | 0.359 | 0.000318 | 0.286 | 0.0519 |
| *CSF1* | 1.030 | 4.14e-12 | -0.165 | 0.00263 | -0.206 | 0.000105 |
| *VIM* | 0.173 | 0.000103 | 0.142 | 0.0058 | 0.283 | 0.000249 |
| *PLAU* | -0.889 | 9.16e-13 | -0.479 | 0.000395 | -0.383 | 0.0181 |
| *GLUL* | 0.366 | 4.1e-12 | 0.029 | 0.495 | -0.092 | 0.0356 |
| *HLA-DRB1* | 0.961 | 0.0315 | 0.458 | 0.426 | -0.374 | 0.481 |
| *HCP5* | -0.969 | 8.84e-07 | -1.299 | 6.13e-06 | -1.196 | 0.00026 |
| *CD274* | 3.554 | 1.42e-14 | -0.741 | 3.9e-08 | -0.659 | 7.79e-05 |
| *HSD3B7* | 1.275 | 6.01e-13 | 0.010 | 0.875 | -0.031 | 0.692 |
| *MYL3* | -0.611 | 0.0128 | -0.671 | 0.00479 | -0.278 | 0.383 |
| *MYH7* | 1.160 | 4.49e-09 | 0.056 | 0.72 | 0.222 | 0.246 |
| *ADAMTSL1* | -0.099 | 0.0238 | -0.407 | 2.22e-06 | -0.132 | 0.101 |
| *SBK2* | 0.376 | 0.0632 | 0.197 | 0.297 | 0.660 | 0.0222 |
| *SLC2A5* | 4.315 | 1.35e-17 | 0.702 | 0.000315 | 0.623 | 0.000802 |
| *ACTA1* | 0.525 | 0.0621 | 0.181 | 0.34 | 0.700 | 0.0784 |
